# Factors that influenced testing positive and dying from COVID-19 in the West South-Central Division of the United States and its Effects on Future Public Health Policy

**DOI:** 10.1101/2025.08.01.25332736

**Authors:** Aloyce R Kaliba

**Affiliations:** Department of Accounting, Finance & Economics, College of Business, Southern University and A&M College, Baton Rouge, LA, USA

**Keywords:** Dying, COVID-19, Generalized Additive Models, Spatial Effects, Testing Positive

## Abstract

**Objectives:** Examine the relationship between various demographic characteristics and meso variables measuring social vulnerability, religiosity, political partisanship, and the built environment on the probability of testing positive for COVID-19 and dying after testing positive for the virus.

**Methods:** The individual-level variables are from the data collected by the Louisiana Department of Health and Hospitals. Meso variables were sourced from various platforms. The data are analyzed using a spatial bivariate probit model with copulas in a generalized additive model framework.

**Results:** The main results suggest a strong and positive association between individual-level covariates and testing positive for the virus and dying from it. The effects of social vulnerability, religiosity, political partisanship, and the built environment varied non-linearly; their effects were within a given critical range.

**Conclusions:** To mitigate the impact of future pandemics like COVID-19, public health policies should focus on addressing existing health disparities, fostering meaningful engagement with community institutions and diverse leaders, and applying proven and scientific public health considerations, while minimizing the influence of political ideologies and culture.

## INTRODUCTION

Following the outbreak of severe acute respiratory syndromeCOVID-19 2 (SARS-CoV-2), also known as COVID-19, in 2019, the World Health Organization declared a global pandemic on March 11, 2020. Subsequently, it removed it from its list of public health emergencies of international concern on May 5, 2023. The availability of multiple COVID-19 vaccines has significantly impacted public health by reducing severe illness and hospitalizations, averting millions of cases and deaths. For example, in the U.S., in 2021, there were about 5,000 reported deaths per day, which receded to less than 300 per week in 2024. However, the Centers for Disease Control and Prevention (CDC) still recommends that everyone 6 months and older receive a COVID-19 vaccine. Recent reports indicate that COVID-19 infection rates surged in the summer of 2024 compared to the summer of 2022 and 2023. Reports on persistent COVID-19 symptoms are also on the rise, as millions of Americans reported long COVID-19 symptoms in 2024 [1] due to the Omicron 2 variant of SARS-CoV-2, and the death rate was about 350 per week from February to June 2025.

After the outbreak, various studies analyzed the determinants of COVID-19 diffusion at the regional level; see, for example, [2,3,4]. While state and country-level studies have broader public health policy implications, individual-level analysis locates the cause of the health outcome at the individual level, within their immediate circle, or among human actors. Individual-level analysis helps explain a person’s traits or behaviors [2], which is vital in identifying the characteristics of the human decision-making process and group health outcomes. However, the extant literature on COVID-19 lacks studies that use extensive data and jointly analyze factors associated with testing positive and subsequent COVID-19-related deaths. The available studies use small samples and focus on two aspects: testing positive or death. To mention a few, see [5,6] for a list of studies, and [7,8] for case studies. Apart from the small sample and lack of joint estimation, the limitations of these studies are twofold: the analysis cannot provide a nuanced understanding of the relationships between the two outcomes (i.e., testing positive or dying), and the interaction of spatial (space) and temporal (time) variables.

This study utilizes patient-level data, including age, gender, and race, in conjunction with meso- and built-environment variables at the U.S. Census Zip Code Tabulation Areas (ZCTAs) and County levels to estimate the probability of testing positive and dying if an individual contracts the virus. The study has two research questions. Did an individual diagnosed with COVID-19 die? The answer is yes or no. If yes, what were the primary sociodemographic and meso-level contributing factors? Therefore, the study has two alternative hypotheses. First, individual and meso-level variables predisposed the SARS-CoV-2 diffusion rate, influencing the probability of COVID-19-related deaths at the personal level. Second, it is essential to note that patients who participated in COVID-19 testing reside in specific areas. Econometrically, a null hypothesis is that the observed health outcomes are spatially random, meaning no systematic spatial relationship exists. Also, since COVID-19-related deaths are observable only after testing positive, those who die might not be representative of the population who participated in the testing exercise. Because the dependent variables are binary (i.e., the patient tested positive: yes or no, if tested positive, died or not), the data were analyzed using a bivariate probit model with a non-random model selection under joint unobserved utility components popularized by Heckman [9] and applied by Marra & Radice [10, 11] for joint estimation.

The data source for this study is the Louisiana Department of Health and Hospitals, available to the author upon special request. For each individual who participated in COVID-19 testing, the Department collected patient demographic data, including age, gender, race, and zip code, which were cross-referable to the U.S. Bureau of the Census’s Zip Code Tabulation Areas (ZCTAs). Meso-level socioeconomic and demographic variables at the ZCTA and county levels are from the 2022-2027 five-year community survey. Additional variables from various sources were measured to assess social vulnerability and political beliefs, accounting for ideological opposition to the SARS-CoV-2 vaccine. Other variables measured religiosity, vaccine supply and demand-related challenges, and environmental and built-environment variables that might increase or reduce the probability of contracting the virus or dying. Due to its advantages, various disciplines, such as strategic management [12] and law [13], among others, apply the bivarite model primarily for jointly determining the influence of covariates on two binary outcomes. For this study, the two equations estimate the individual’s risk of testing positive for COVID-19 and their risk of dying once infected.

The main contributions of the study are twofold: (i) add to the COVID-19 literature regarding how individual, meso, and built-environmental variables influence testing positive and dying, and (ii) instead of estimating a single linear parameter for each covariate, as Baayen & Linke (2020) show, with the generalized additive models it is possible to display how the slope of a response variable changes across the range of a covariate. Moreover, it is crucial to distinguish this study from other population studies, as it focuses on a sample that has truly participated in the COVID-19 testing exercise and might not be fully representative of the population. The study’s results are helpful for the surveillance of infectious diseases, the control of future outbreaks, supporting vaccination campaigns for various diseases, and informing future research. The results are also significant when implementing sustainable and impactful public health measures that use theoretically informed approaches to guide the design, development, implementation, and evaluation of public health interventions. Such an understanding is crucial for effective policymaking, particularly when interacting with various aspects of individuals and communities, as well as for planning considerations that promote sustainable and healthy futures. Additionally, Walugembe et al. [14] suggest that appreciating the effects of individual demographics on health outcomes enables long-term improvements in public health by maximizing the limited available resources through targeted community support.

## METHODS

### Sample and Source of Data

Figure 1 illustrates the location of individual patients at the ZCTA level in Louisiana and ZCTAs in Arkansas, Texas, and Oklahoma. In Figure 1, the West South Central Division includes Arkansas, Oklahoma, Louisiana, and Texas. Based on the sample point distribution, the study area comprises twelve regions, including Arkansas, Oklahoma, Texas, and eight regions in Louisiana. Louisiana has eight regions that comprise the Louisiana Association of Planning and Development Districts (LAPDD), a network of multi-service organizations, including the Acadian, Bayou, Capital Region, Central, Northeast, Northwest, Southeast, and Southwest regions, as described on their website at https://lapdd.org/. The objective of the LAPDD is to foster cooperation and collaboration among local governments and other stakeholders to address regional planning and development challenges in Louisiana.

**Figure 1.**
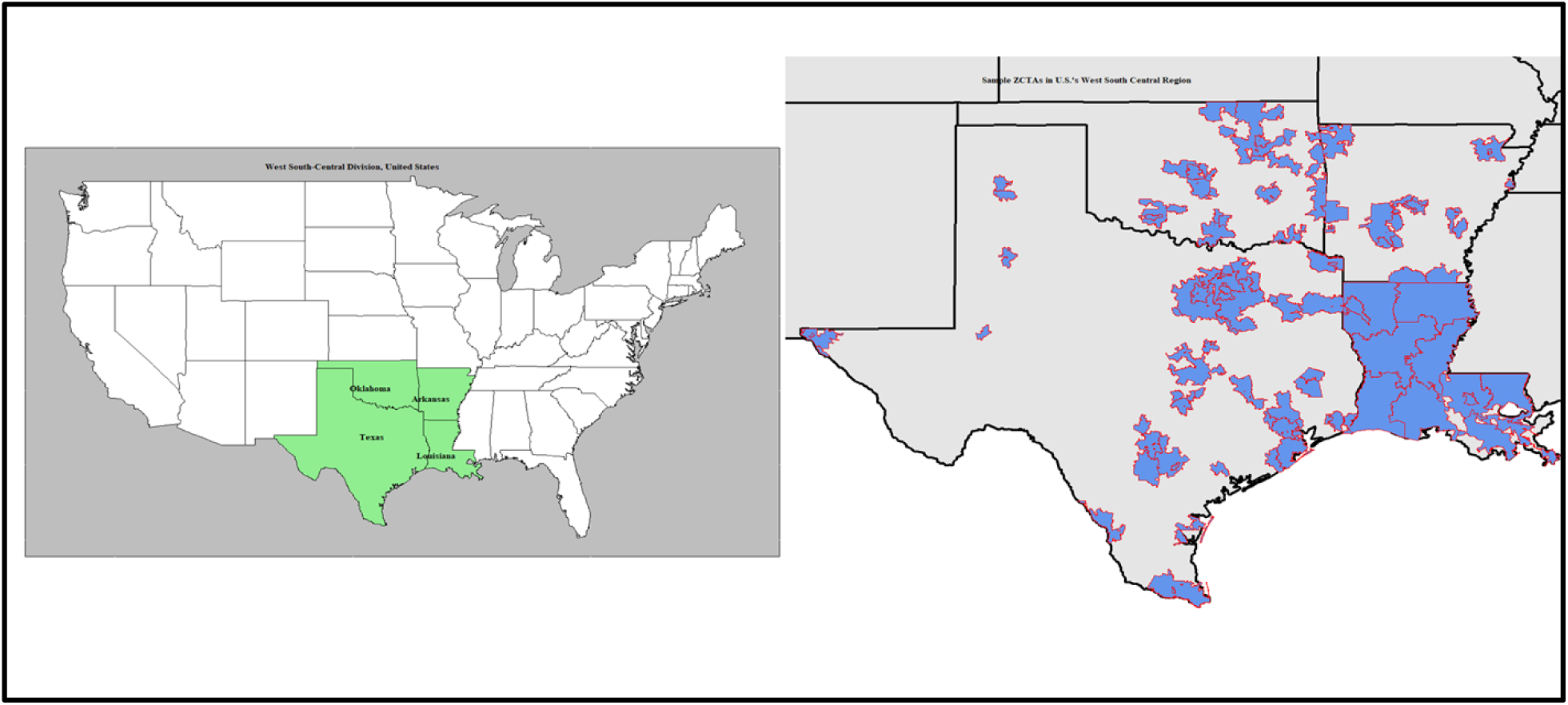
Location of Observed Samples in Some Zip Code Tabulated Areas in the West South-Central Division, United States.

The individual-level data are from the Louisiana Department of Health and Hospitals, obtained through a special request. The data were cross-matched using the ZIP Code and ZCTA geoids. The individual-level data is from March 1, 2020, to October 1, 2022. For each reported COVID-19 case, the patient has a unique identification number and demographic characteristics, including age, gender, race, zip code, and the date the patient tested positive or died. Meso-level variables were from various Geographical Information Systems and Platforms. The data includes social vulnerability, vaccine distribution challenges, and political and religious beliefs. Other meso variables measured included measures of congestion and pollution, as well as the community-level built environment. Based on availability, these variables were at the ZCTAs, census tract, or county level. The data were cross-matched using the ZIP Code and ZCTA geoids.

The Centers for Disease Control and Prevention (CDC) provides data on the Social Vulnerability Index (SVI) at https://www.atsdr.cdc.gov/place-health/php/svi/index.html. The index is at the county level and aggregates demographic and socioeconomic characteristics to measure the relative social vulnerability of U.S. communities to various hazards, including natural disasters, disease outbreaks, and environmental risks. It combines socioeconomic, demographic, housing, and transportation factors that can exacerbate the impacts of these events. The 2020 SVI ranges from 0 to 1, with a higher value indicating greater social vulnerability [15]. The vaccine distribution challenge is measured using the Surgo COVID-19 Vaccine Coverage Index for February 2021 at the country level. The index catches the supply- and demand-related challenges hindering the rapid spread of SARS-CoV-2 vaccine coverage. It measures the level of concern for a problematic rollout from 0 (no significant problem) to 1 (considerable severe problem) with five categories: very-low concern (score of 0–0.2); low concern (0.2–0.4); moderate concern (0.4–0.6); high concern (0.6–0.8), and very-high concern (0.8–1). The index represents five specific subthemes: historic under-vaccination, sociodemographic barriers to vaccination, resource-constrained healthcare systems, healthcare accessibility barriers, and irregular care-seeking behaviors [16], available at https://vaccine.precisionforcovid.org/.

The Cook’s Partisan Voter Index, as explained at https://www.cookpolitical.com/cook-pvi, captures the ideological dimensions at the county level. The Cook Partisan Voting Index (PVI) measures how strongly a U.S. congressional district or state leans towards either the Republican or Democratic party at the presidential level, compared to the nation as a whole. It is a numerical score that helps assess the partisan landscape and competitiveness of districts or states. It measures how strongly a county has leaned toward the Democratic or Republican Party for the past two Presidential elections. A positive value in the Cook Partisan Voting Index (PVI) indicates that a particular congressional district or state leans more Republican compared to the national average. Negative PVI values are associated with Democratic-leaning states or districts. The data used to calculate the 2020 PVI are the results of the 2016 and 2020 presidential elections, sourced from the Massachusetts Institute of Technology (MIT) Election Data and Science Lab at https://electionlab.mit.edu/data.

Additionally, the Statisticians of American Religious Bodies report adherence rates per 1,000 people as a measure of religiosity. The adherence rate indicates the number of people affiliated with a particular religious organization, including Evangelical Protestants, Historically Black Protestants, White Mainline (non-Evangelical) Protestants, Catholics, and other major Christian groups in the United States. Based on the 2020 census data collected by the Association of Religion, the data is accessible from https://www.usreligioncensus.org/maps and https://thearda.com/data-archive/browse-categories#. The data provide information on church membership and average weekly attendance at worship services, enabling the estimation of the religious adherence rate by denomination within the U.S. population and by county.

Regarding pollution, the Environmental Protection Agency (EPA) reports country-level air quality indices. The Air Quality Index (AQI) is a system used to communicate to the public the level of air pollution and its potential health effects. It is based on the levels of specific air pollutants, calculated using measurements or forecasts of these pollutants. The AQI scale typically ranges from 0 to 500, with higher numbers indicating more polluted air and greater health risks. The main components of the Air Quality Index (AQI) are six common air pollutants: ozone, particulate matter (PM2.5 and PM10), carbon monoxide, sulfur dioxide, and nitrogen dioxide. The data is available at https://www.epa.gov/aqs/obtaining-aqs-data. When the AQI is above 150, it is unhealthy for everybody. Unhealthy days, therefore, refer to the number of days in a year when the index was above 150.

The U.S. Department of Health and Human Services also reports Population-Level Analysis and Community Estimates (PLACES) of the burden of diseases and the geographic distribution of health outcomes. PLACES, or “Local Data for Better Health,” is a collaboration between the Centers for Disease Control and Prevention (CDC), the Robert Wood Johnson Foundation, and the CDC Foundation. It provides health data for small areas across the United States, including counties, places, census tracts, and ZIP Code Tabulation Areas (ZCTAs). The data is downloadable from https://www.cdc.gov/places/about/index.html. The PLACES data helps local health departments and jurisdictions understand the burden and geographic distribution of health measures, enabling them to plan effective public health interventions. The compilation of PLACES data is by merging data from the Behavioral Risk Factor Surveillance System (BRFSS) and the American Community Survey. The data cover a range of topics, including health outcomes (e.g., the proportion of the population who are obese or diabetic), preventive service use, health risk behaviors, disabilities, health status, and social determinants of health.

The built-environment variables are from the U.S. EPA’s Smart Location Database available at https://www.epa.gov/smartgrowth/smart-location-mapping. The 2021 database presents indicators related to the built environment and location efficiency of various areas in the United States. These indicators measure the density of development, diversity of land use, street network design, and accessibility to destinations, as well as various demographic and employment statistics. Notably, the Smart Location Score (SLC) or Smart Location Index (SLI) represents the location efficiency of residential areas. The EPA SLC is a measure of how well a location supports sustainable transportation and land use patterns. Based on the Smart Location Database, the index aggregates population density, land use diversity, built environment design, access to destinations, and proximity to transit. The score helps evaluate the potential for reduced vehicle miles traveled and greenhouse gas emissions associated with a particular location. The SLC ranges from 0 to 100; a higher score indicates a more location-efficient site (reduced congestion and pollution), and a lower score indicates a more location-inefficient site (increased congestion and pollution).

#### Variable Selection and Empirical Model

The available literature guided the selection of variables for this study. Refer to Appendix 1 for the list of variables included in the global baseline model. Two considerations were applied when estimating the probability of testing positive and dying from the virus. First, past studies have identified individual-level covariates, meso-level variables, community-built environments, and environmental factors that influence the likelihood of testing positive for COVID-19 or dying from the virus after a positive test. For example, see [16, 17, 18, 19], among many others. Reducing the number of redundant covariates and simplifying the model can be achieved by selecting the best predictor subsets using a backward variable selection method, as described by Sutter & Kalivas [20] and implemented in the H2O package by Fryda et al.[21] in the R Software environment [22]. Logistic regression was applied to each Equation (i.e., testing positive or dying) to identify redundant variables. See Appendix 1 for variables included in the complete model.

Second, after selecting relevant variables, a spatial bivariate model, explained by [10, 23], was estimated to account for both non-random selection and spatial effects. The model addressed selection bias by incorporating a selection equation (testing positive) alongside the outcome equation (dying from the virus), while controlling for the possibility that the observed sample is not representative of the population. Since individuals reside in predetermined neighborhoods, according to Manski [24], the relationship between the response variable and the covariates can vary by location (spatial heterogeneity), covariates in one region might correlate with similar variables in other places (spatial autocorrelation), and similar unobserved characteristics among locations result in spatially correlated effects in the model’s error term [25]. To account for spatial relationships, the standard approach uses distance-based and Kernel-based matrices to capture the spatial relationships in the data. These weights are complex to implement, interpret, and validate, and are generally computationally intensive, especially for large datasets.

The generalized additive models [26] establish spatial relationships between covariates and the response variable through smooth patterns that can be linear or non-linear using the location geocode. Marra & Wood [27] present an overview of smooth selection and hypothesis testing approaches for determining the appropriate smooths for each covariate, utilizing prediction error criteria or likelihood-based methods. Generalized additive models (GAMs) incorporate smooth terms that capture non-linear relationships between predictors and the response variable using spatial coordinates (i.e., longitudes and latitudes) within the model. This approach enables spatially varying coefficients, thereby reducing spatial heterogeneity and capturing the data’s underlying spatial structure and trends, effectively removing or minimizing the effects of autocorrelation. Econometrically, the estimated complete model is as follows:

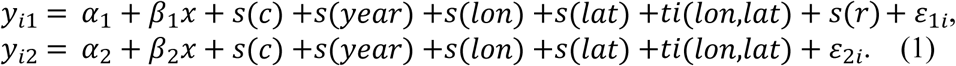

In Equation (1), the dependent variables (*y*_*i*1_) and (*y*_*i*2_) are binary and equal to one for those who tested positive or died after testing positive and zero otherwise for the ith observation, respectively. The covariates (*x*) and (*c*) are a mix of factors and continuous variables at the individual and meso-levels, respectively, where *s(.)* is a smooth function. Other covariates include the year (i.e., 2019, 2020, and 2021). Lon and lat represent the ZCTA’s centroid (i.e., longitude and latitude), used as location variables. The covariate (*r*) is a factor variable representing regions in Figure 1, and it accounts for exclusion restriction [28], implying that the regions’ influence is on testing positive but not dying from the virus.

Note that the smooths of covariates (*year*) and (*r*) were declared as a random effect basis (bs = “re”) to include time and regional random effects. Argument *bs* specifies the type of spline basis employed, which is *re* (random effect). For continuous variables, the smooth was such that *s(c, bs=“ts”)*, specifying that the smooth term for continuous variable *c* should be a shrinkage thin plate regression spline. Shrinkage thin plate regression splines offer several advantages, particularly in the presence of noisy or high-dimensional data. They avoid the “knot placement” issues associated with conventional regression splines by using all input points as knots and then shrinking the coefficients through regularization, thereby providing a more flexible and robust approach to estimating smooth functions [29].

The *ti(.)* function in Equation (1) creates a smooth tensor product by combining the effects of separate smooth functions for individual variables and their interactions, particularly when they are on the same scale. The implied *ti(lon, lat, bs=“tp”, k=50)* function creates a spatial smooth that captures how the effect of covariates changes across ZCTAs, which achieves a full rank [30]. The spatial smooth term is defined using “tp,” creating a thin-plate regression spline, a common choice for spatial data, and applying it to longitude and latitude as a single tensor product smooth term. The basis dimension (*k*) should be large enough to capture the full spatial complexity. The two Equations were jointly estimated using the GJRM package [11] as a bivariate probit model within the R software environment [22]. It is important to note that the probit link function transforms the probability of the outcome into a z-score (the inverse of the cumulative normal distribution function). The transformation is linear, implying that changes in the linear predictor directly reflect changes to the z-score, or a corresponding shift in the probability distribution. To interpret the results in terms of probabilities, you need to back-transform the predicted z-scores (z) using the 1-Φ(z) formula, where Φ(.) is the cumulative normal distribution function. As Baayen & Linke [31] show, the plot of the smooth for *ti(.)* displays how the slope of a fitted function changes across the range of a covariate.

The results of the complete model are compared to those of the restricted model, which excludes spatial effects, using the Bivariate Normal (N) or Joe 90 (J90) copulas. Nelsen (2006) and Durante & Sempi (2016), A copula is a multivariate cumulative distribution function (CDF) with uniform marginal distributions ranging from 0 to 1 [32]. It describes the dependence structure between random variables, allowing for the modeling of correlations and other forms of association, even when the individual variables have non-uniform distributions [33]. The copulas construct joint distributions by combining uniform marginals and capturing the dependence structure separately from the marginal behavior. A bivariate normal copula is a special type that models the joint distribution of two random variables where the margins are uniform between 0 and 1 [34]. A bivariate normal copula uses the cumulative distribution function of the bivariate standard normal distribution with a specified correlation parameter.

As Joe, Li & Nikoloulopoulos [35] show, the Joe copula models dependence between two or more random variables. The copula is well-suited for situations with moderate to strong positive dependence and moderate to heavy tail dependence. It offers flexibility in modeling various dependence structures not captured by simple correlation coefficients. Particularly, the Joe copula with a 90-degree rotation [36] is helpful when the dependence between response variables is not linear or does not follow a simple elliptical structure. Specifically, the Joe-90 copula is applied when response variables have tail asymmetry or tail dependence, implying that the extreme values (tails) of the distributions of the response variables might show a stronger relationship than the rest of the data. The copula is convenient when multivariate normality is not a reasonable assumption. Specifically, the Joe-90 copula is a versatile tool for modeling dependence structures, notably when the data deviates from the assumptions of simpler models, and in scenarios where one variable experiences extreme losses (i.e., a higher probability of dying). At the same time, the other exhibits extreme gains (higher probability of testing positive), as individuals who test positive may have a higher risk of death from the virus.

## RESULTS AND DISCUSSION

### Distribution of Response and Individual-Level Variables

The results in Table 1 present COVID-19 test results, deaths, and the distribution of unique individuals based on the sample data. The dataset comprised 18,204,894 observations, representing actual COVID-19 tests. Of these tests, 18,057,962 were from Louisiana, 130,278 from Texas, and 8,898 and 7,756 from Oklahoma and Arkansas, respectively. The data is from March 1, 2020, to January 21, 2022. In 2020, during the experimental management period, approximately 6.12 million COVID-19 tests were conducted in Louisiana, involving 1.75 million unique individuals. For Louisiana in 2020. 5.49 million tests were negative, 618,240 test results were positive, and 8,953 test results were inconclusive. The recorded test results in 2020 by the Louisiana Department of Health and Hospitals for the region were approximately 6.15 million, with 89.77% of the tests yielding negative and 10.09% yielding positive results. In 2020, about 59,512 patients in Louisiana died due to the virus. The results for 2021 (the lockdown period with the alpha and delta variants) and 2022 (the delta vaccine period with the Omicron variant) have a similar interpretation. The year with the highest positive test results was 2022, at 28.87 percent, while the majority of deaths occurred in 2020, at 62.96 percent. Comparatively, the CDC data summarized by Ahmad [37] indicate that in 2020, there were approximately 384,536 (35.25%) deaths in the United States attributed to COVID-19, compared to 462,193 (42.37%) and 244,000 (22.37%) COVID-19 deaths in 2021 and 2022, respectively.

**Table 1.** COVID-19 test results, number of unique individuals, and deaths from the sample.

| **Year** | **Test Results** | **Arkansas** | **Louisiana** | **Oklohama** | **Texas** | **Total** | **Percentage** |
| --- | --- | --- | --- | --- | --- | --- | --- |
| **COVID-19 test results** |  |  |  |  |  |  |  |
| 2020 | Inconclusive | 5 | 8,953 | 0 | 24 | 8,982 | 0.15% |
|  | Negative | 2,708 | 5,488,027 | 767 | 32,065 | 5,523,567 | 89.77% |
|  | Positive | 288 | 618,240 | 66 | 2,113 | 620,707 | 10.09% |
|  | Sub_Total | 3,001 | 6,115,220 | 833 | 34,202 | 6,153,256 | 100.00% |
| 2021 | Inconclusive | 1 | 13,692 | 155 | 74 | 13,922 | 0.13% |
|  | Negative | 4,025 | 9,598,781 | 6,877 | 85,771 | 9,695,454 | 91.02% |
|  | Positive | 310 | 938,075 | 938 | 3,890 | 943,213 | 8.85% |
|  | Sub Total | 4,336 | 10,550,548 | 7,970 | 89,735 | 10,652,589 | 100.00% |
| 2022 | Inconclusive | 2 | 3,926 | 0 | 11 | 3,939 | 0.28% |
|  | Negative | 298 | 986,041 | 72 | 4,802 | 991,213 | 70.85% |
|  | Positive | 119 | 402,227 | 23 | 1,528 | 403,897 | 28.87% |
|  | Sub Total | 419 | 1,392,194 | 95 | 6,341 | 1,399,049 | 100.00% |
| Grand Total | Inconclusive | 8 | 26,571 | 155 | 109 | 26,843 | 0.15% |
|  | Negative | 7,031 | 16,072,849 | 7,716 | 122,638 | 16,210,234 | 89.04% |
|  | Positive | 717 | 1,958,542 | 1,027 | 7,531 | 1,967,817 | 10.81% |
|  | Total | 7,756 | 18,057,962 | 8,898 | 130,278 | 18,204,894 | 100.00% |
| **Number of Unique individuals** |  |  |  |  |  |  |  |
| **Year** |  | **Arkansas** | **Louisiana** | **Oklohama** | **Texas** | **Total** | **Percentage** |
| 2020 |  | 1,004 | 1,749,923 | 317 | 13,002 | 1,764,246 | 35.36% |
| 2021 |  | 1,385 | 2,511,841 | 3,022 | 23,070 | 2,539,318 | 50.90% |
| 2022 |  | 201 | 681,597 | 56 | 3,679 | 685,533 | 13.74% |
| Grand Total |  | 2,590 | 4,943,361 | 3,395 | 39,751 | 4,989,097 | 100.00% |
| **Dying due to the COVID-19 Virus** |  |  |  |  |  | **0** |  |
| **Year** |  | **Arkansas** | **Louisiana** | **Oklohama** | **Texas** | **Total** | **Percentage** |
| 2020 |  | 14 | 59,512 | 2 | 53 | 59,581 | 62.96% |
| 2021 |  | 13 | 32,408 | 0 | 70 | 32,491 | 34.33% |
| 2022 |  | 0 | 2,560 | 0 | 4 | 2,564 | 2.71% |
| Grand Total |  | 27 | 94,480 | 2 | 127 | 94,636 | 100.00% |

Tables 2 through 4 present the distribution of individual-level variables based on the number of COVID-19 test results. Note that the grant total in Table 2 (18,166,006 tests) and Table 4 (18,198,848) differs slightly from that in Table 1 (18,204,894 tests) due to missing values in the gender and age-group variables. The grant total in Table 3 is similar to that in Table 1. The results in Table 2 show that, in all three years, the majority of participants (61.34%) in COVID-19 testing were female. Women are more likely to perceive COVID-19 as a severe health problem, to agree with preventive public policies, and comply with restrictive measures, including regular testing [38, 39, 40].

**Table 2.** Sample distribution by gender.

| **Year** | **Gender** | **AR** | **LA** | **OK** | **TX** | **Total** | **Percentage** |
| --- | --- | --- | --- | --- | --- | --- | --- |
| 2020 | Female | 1,702 | 3,782,874 | 280 | 16,544 | 3,801,400 | 61.78% |
|  | Male | 1,282 | 2,316,404 | 553 | 17,546 | 2,335,785 | 37.96% |
|  | Unidentified | 17 | 15,942 | 0 | 112 | 16,071 | 0.26% |
|  | Sub-total | 3,001 | 6,115,220 | 833 | 34,202 | 6,153,256 | 100.00% |
| 2021 | Female | 2,273 | 6,438,222 | 1,431 | 44,429 | 6,486,355 | 61.11% |
|  | Male | 2,048 | 4,073,664 | 6,529 | 45,090 | 4,127,331 | 38.89% |
|  | Unidentified | 15 |  |  |  | 15 | 0.00% |
|  | Sub-total | 4,336 | 10,511,886 | 7,960 | 89,519 | 10,613,701 | 100.00% |
| 2022 | Female | 215 | 853,172 | 27 | 2,615 | 856,029 | 61.19% |
|  | Male | 202 | 534,340 | 64 | 3,692 | 538,298 | 38.48% |
|  | Unidentified | 2 | 4,682 | 4 | 34 | 4,722 | 0.34% |
|  | Sub-total | 419 | 1,392,194 | 95 | 6,341 | 1,399,049 | 100.00% |
| Grand total | Female | 4,190 | 11,074,268 | 1,738 | 63,588 | 11,143,784 | 61.34% |
|  | Male | 3,532 | 6,924,408 | 7,146 | 66,328 | 7,001,414 | 38.54% |
|  | Unidentified | 34 | 20,624 | 4 | 146 | 20,808 | 0.11% |
|  | total | 7,756 | 18,019,300 | 8,888 | 130,062 | 18,166,006 | 100.00% |

**Table 3.** Sample Distribution by Race.

| Year | Race Group | AR | LA | OK | TX | Total | Percentage |
| --- | --- | --- | --- | --- | --- | --- | --- |
| 2020 | African American/black | 1,085 | 2,349,416 | 158 | 9,154 | 2,359,813 | 38.35% |
|  | Asian | 29 | 157,388 | 32 | 647 | 158,096 | 2.57% |
|  | White | 1,135 | 2,692,547 | 323 | 13,813 | 2,707,818 | 44.01% |
|  | Others | 752 | 915,869 | 320 | 10,588 | 927,529 | 15.07% |
|  | Sub-Total | 3,001 | 6,115,220 | 833 | 34,202 | 6,153,256 | 100.00% |
| 2021 | African American/black | 1,340 | 4,347,745 | 1,388 | 26,560 | 4,377,033 | 41.09% |
|  | Asian | 67 | 195,174 | 128 | 1,922 | 197,291 | 1.85% |
|  | White | 2,361 | 4,574,394 | 4,308 | 35,889 | 4,616,952 | 43.34% |
|  | Others | 568 | 1,433,235 | 2,146 | 25,364 | 1,461,313 | 13.72% |
|  | Sub-Total | 4,336 | 10,550,548 | 7,970 | 89,735 | 10,652,589 | 100.00% |
| 2022 | African American/black | 115 | 587,099 | 24 | 1,682 | 588,920 | 42.09% |
|  | Asian | 2 | 29,614 | 3 | 76 | 29,695 | 2.12% |
|  | White | 212 | 592,742 | 43 | 2,454 | 595,451 | 42.56% |
|  | Others | 90 | 182,739 | 25 | 2,129 | 184,983 | 13.22% |
|  | Sub-Total | 419 | 1,392,194 | 95 | 6,341 | 1,399,049 | 100.00% |
| Grand total | African American/black | 2,540 | 7,284,260 | 1,570 | 37,396 | 7,325,766 | 40.24% |
|  | Asian | 98 | 382,176 | 163 | 2,645 | 385,082 | 2.12% |
|  | White | 3,708 | 7,859,683 | 4,674 | 52,156 | 7,920,221 | 43.51% |
|  | Others | 1,410 | 2,531,843 | 2,491 | 38,081 | 2,573,825 | 14.14% |
|  | Sub-Total | 7,756 | 18,057,962 | 8,898 | 130,278 | 18,204,894 | 100.00% |

**Table 4.**
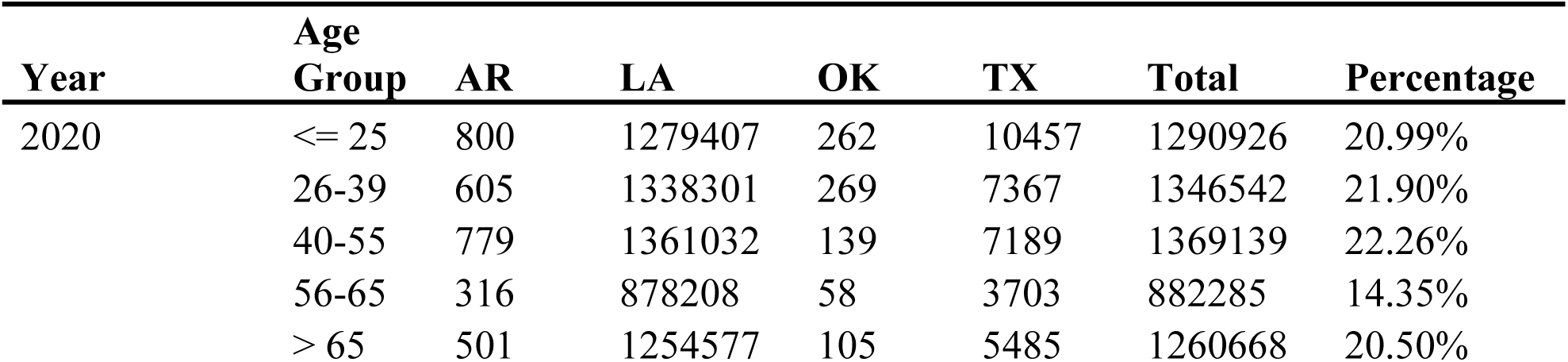
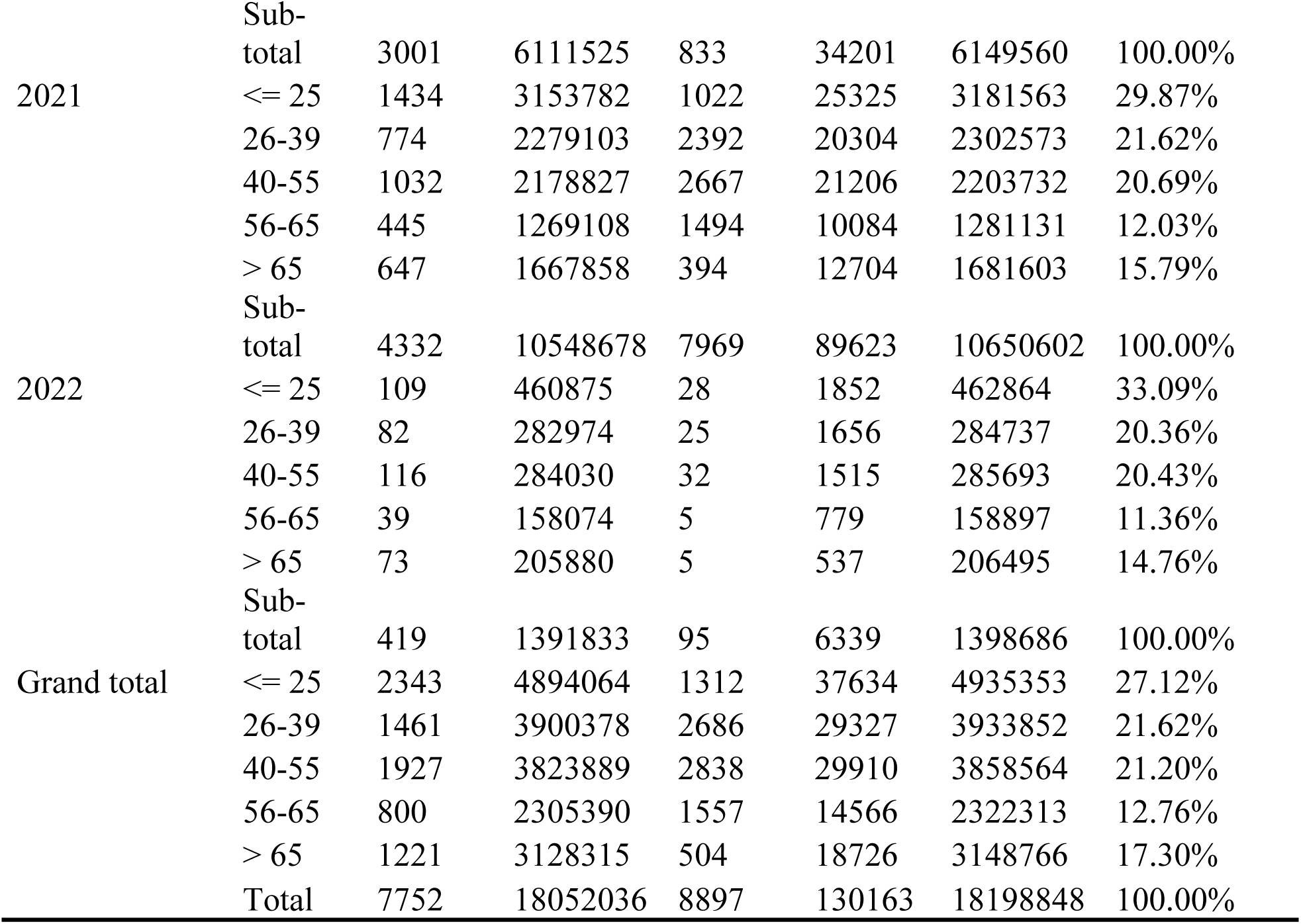
Sample distribution by age.

According to the 2023 American Community Survey year 1 estimates, the population in the West South-Central Division was 42,198,608 people living in 1,100,942 square kilometers, implying a population density of 38 people per square kilometer. The population distribution by race was as follows: White (45%), Hispanic (31%), African American (14%), Asian (5%), and others (5%). The median age was 36.5 years compared to 39.2 years in the United States. The median age of the division was 36.5, compared to 39.2 years for about 90 percent of the United States population. In Table 3, the average participation of white people in COVID-19 testing (43.51%) was close to the percentage of white people in the population. The average participation in COVID-19 testing by black people (40.24%) was relatively high compared to the fraction of black people in the population, which is 14 percent. As Hamilton-Burgess et al. [41] demonstrate, partnerships between safety-net providers and Black community networks facilitated COVID-19 testing outreach activities and increased trust among Black individuals who participated in testing.

The results in Table 4 show that from the sample data, those who participated in COVID-19 testing, about 27.12 percent of the West South Central Division population, fell into the age of 0-25 years category, which increased from 20.99 percent in 2020 to 29.87 and 33.09 percent in 2021 and 2022, respectively. For other age groups, participation in COVID-19 testing was relatively static. The participation rate averaged 21.62 percent for the 26-39 age group, 21.20 percent for the 40– 55 age group, 12.76 percent for the 56– 65 age group, and 17.30 percent for the above-65 age group. Generally, the incidence of COVID-19 increased in all age groups, with the most rapid rate among children and adolescents (aged 0–17 years) and young adults (aged 18–24 years), which might have increased the testing participation rate among the 0-25 age group [42].

### Results of the Generalized Joint Regression Model

Table 5 presents the results from testing the models’ fit for non-spatial and spatial generalized joint regression models with a Bivariate Normal or J-90 Copula, respectively. We use a convergence check function in the GJRM package [11] to determine if the model has converged and provide more information about the score, where the value should be close to zero. The Akaike and Bayesian Information Criteria (AIC & BIC) are traditional statistical tools used for model selection, which often involves comparing multiple candidate models. Both AIC and BIC provide methods for evaluating the quality of a statistical model by balancing goodness of fit and model complexity [27, 29]. The lower the value, the better the model fits the analyzed data.

**Table 5.** Results from testing the models’ fit Non-spatial model with a Bivariate Normal Copula.

|  |  |
| --- | --- |
| **Non-spatial model with a Bivariate Normal Copula** |  |
| Convengence check | 0.00000013 |
| Akaike Information Criterion (AIC) | 2,948,793 |
| Bayesian Information Criterion (BIC) | 2,949,692 |
| **Non-spatial model with J-90 Copula** |  |
| Convengence check | 0.00089471 |
| Akaike Information Criterion (AIC) | 2,278,797 |
| Bayesian Information Criterion (BIC) | 2,289,812 |
| **Spatial model with a Bivariate Normal Copula** |  |
| Convergence check | 0.00000003 |
| Akaike Information Criterion (AIC) | 2,248,395 |
| Bayesian Information Criterion (BIC) | 2,269,939 |
| **Spatial model with copula J-90 Copula** |  |
| Convergence check | 0.00089364 |
| Akaike Information Criterion (AIC) | 2,148,421 |
| Bayesian Information Criterion (BIC) | 2,149,926 |

Note: For all models, the information matrix associated with the fitted model was positive definite.

The results in Table 5 indicate that the convergence scores of the estimated models were close to zero, suggesting that the optimization algorithm successfully converged after finding a set of parameter estimates that maximized the log-likelihood and the gradient of the log-likelihood function (the score) was approximately zero. Additionally, the information matrices associated with all fitted models were positive definite, indicating a well-behaved statistical model with identifiable parameters and a unique, estimable maximum likelihood solution [11]. Positive definiteness ensures that the maximum likelihood estimator is well-defined and that the standard errors of the parameter estimates are accurate [43]. Based on AIC and BIC values, the best model is the Spatial model with the Copula J-90, which has the lowest values. Also, notice that the BIC values are relatively large because the statistic penalizes model complexity more strongly than AIC, especially with larger datasets. Moreover, spatial models performed better than models that did not include spatial effects, as locations closer to each other tend to be more similar, and spatial relationships and dependencies are likely present in the current data. The results from the spatial model with the Copula J-90 are discussed in detail below. For comparison purposes only, the results of the other models are in Appendix 2.

The results of the spatial model with the Copula J-90 are in Table 6. The results are for the first and second equations. The first Equation estimates the probability of testing positive, and the second Equation predicts the likelihood of dying after a positive test result. The last part of the paper shows that, on average, 1.52 percent of those who tested positive died from the range. At a 95 percent confidence level, the range was between 1.48 percent and 1.58 percent. The parametric and smooth estimates are in the first and second parts of the table. For the parametric coefficients, each level of the factor (excluding the reference level) is represented by a coefficient, indicating its deviation from the effect of the reference level on the response variable. For the smooth terms, each smooth has several coefficients - one for each basis function, therefore not provided, and reports the effective degrees of freedom (edf) representing the complexity of the smooth. By definition, the edf of 1 is equivalent to a straight line, the edf of 2 is comparable to a quadratic curve, and higher values describe more complex and wiggly curves. Appendix 3 shows the plot of each smooth in equations 1 and 2.

**Table 6.**
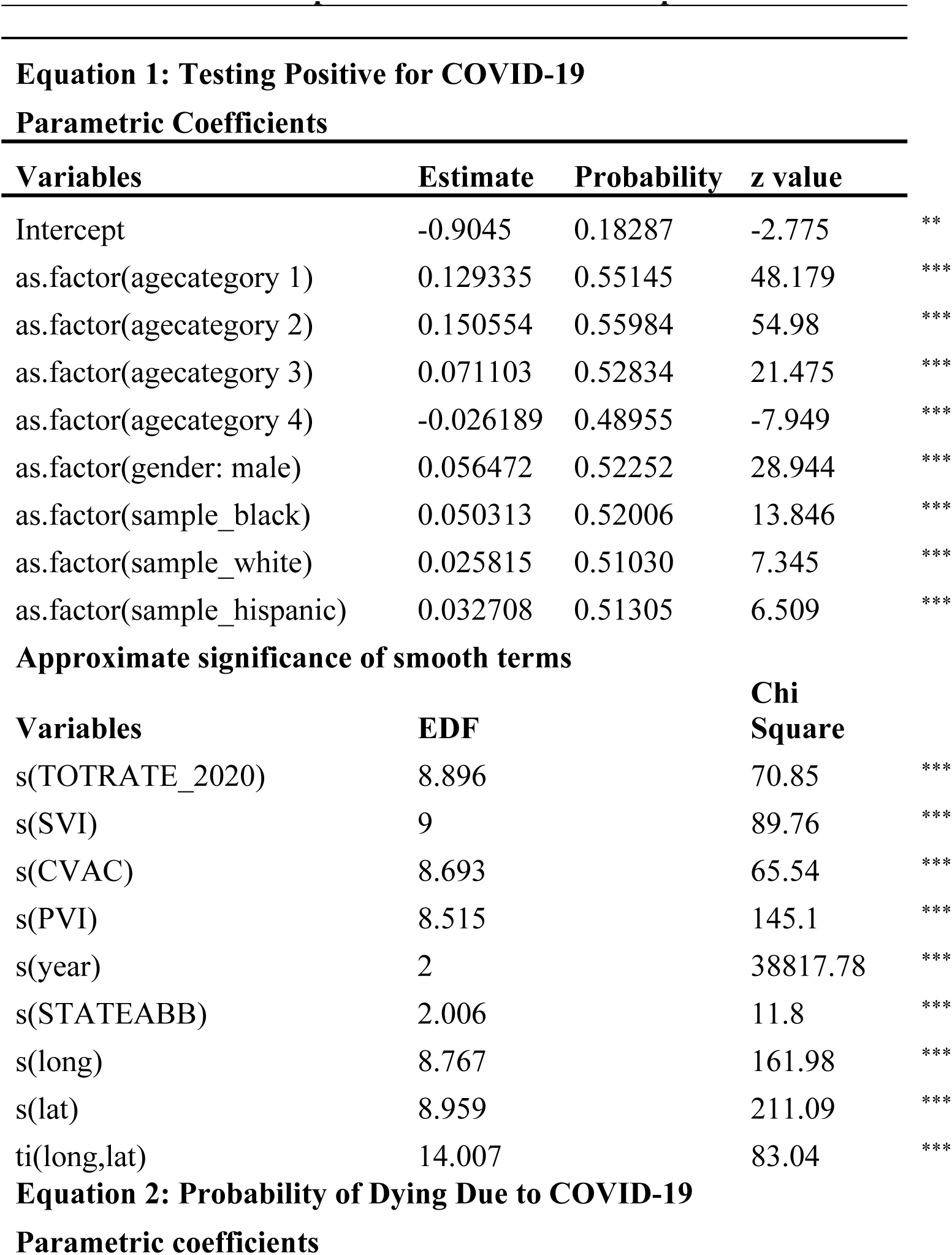
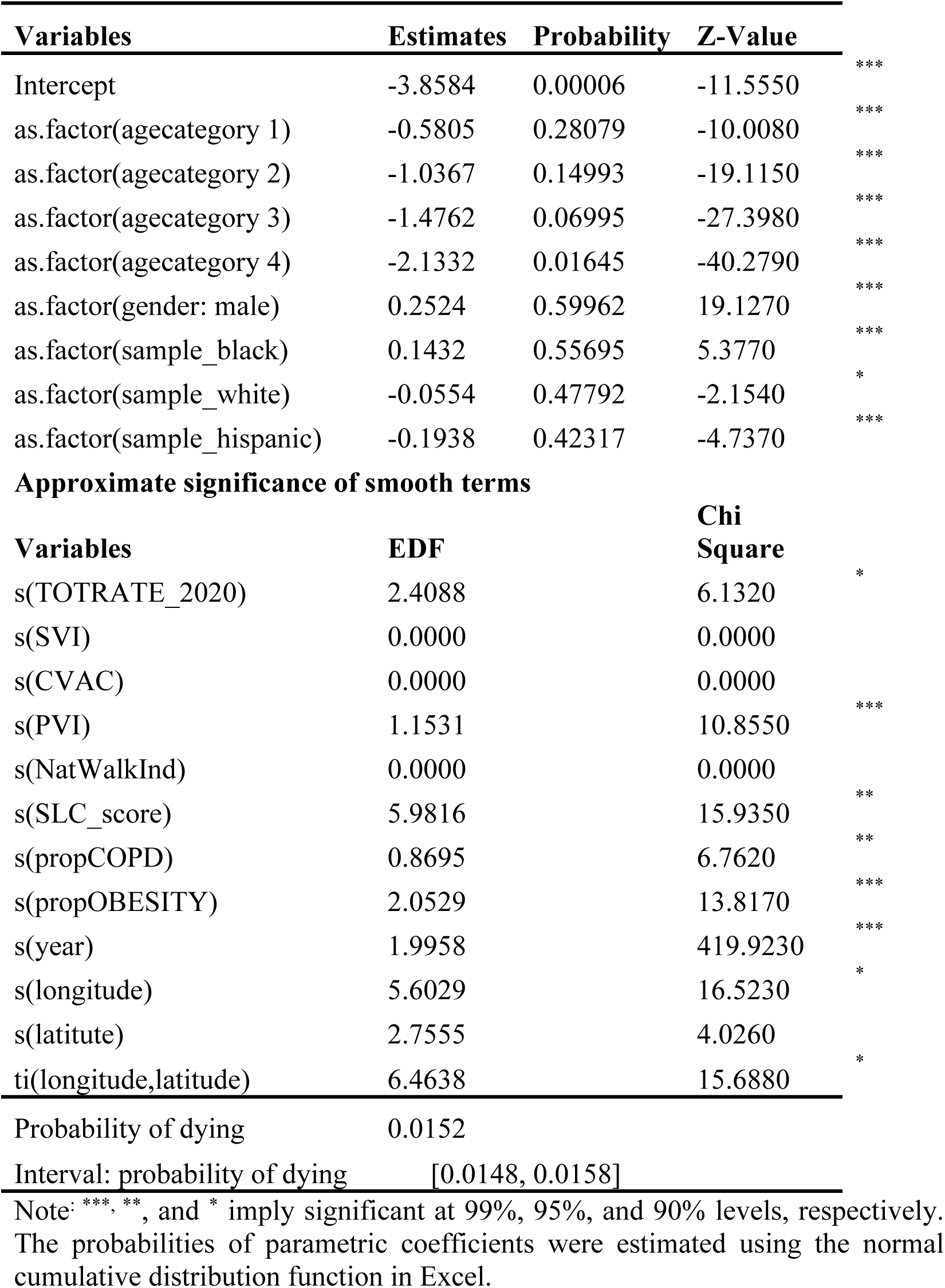
Results of the spatial model with J-90 Copula.

Starting with the parametric coefficients in Table 6, all individual-level variables included in the model (as factors) are statistically significant at a confidence level of less than 0.01. The results suggest a strong real effect between testing positive and individual-level covariates. The estimated probability in Table 6 indicates how much, on average, the likelihood of testing positive or dying increases or decreases from the reference group. In Table 6, the age-group reference category is the group of individuals who are 65 years or older (see Table 4 above). Except for the probability of the 56-65 year age category, which is negative, the coefficients of the other age categories are positive. Being in the 56-65 years age category decreased the probability of testing positive by 2.62 percent compared to those who were older than 65 years. For other age categories, the probability of testing positive increased by 15.10%, 12.93%, and 7.11% for individuals in the 26-39, under 25, and 40-55 age groups, respectively. Except for the 56-65 age group, the probability of testing positive increases with age when compared to individuals older than 65. The trend is attributable to the higher propensity of younger adults to dine out, increased testing, and the rejection of social distancing and mask-wearing [44, 45].

The gender and race variables in Table 6 are dummy variables. For gender, gender=1 if male, zero otherwise, where the reference is female. For race, there are three dummy variables: black=1 if black, otherwise; white=1 if white, otherwise; and hispanic=1 if hispanic, zero otherwise. The reference for these dummies is other races. Males, compared to females, were likely to test positive by 52.25 percent. Other studies have found gender differences in testing positive for COVID-19, with a higher rate among males, accounting for 59–68% of the positive cases [46, 47]. After controlling for socioeconomic and demographic characteristics, the gap is due to differences in biological systems [48, 49].

Regarding race, Black people, compared to other races, were likely to test positive by 52.01 percent. The likelihood of testing positive for White and Hispanic people was high by 51.03 and 51.31percent, respectively, compared to other races. Note that “other races” in America include Asian, American Indian or Alaska Native, and Native Hawaiian or Other Pacific Islander. Therefore, other races for the Black race include White, Hispanic, Asian, American Indian or Alaska Native, and Native Hawaiian or other Pacific Islander. The interpretation is analogous to the White and Hispanic dummy variables. These results do not present a clear picture. Research shows that in Southern states, particularly during the early stages of the COVID-19 pandemic, racial and ethnic minorities, specifically Black and Hispanic populations, were more likely to test positive for COVID-19 compared to White individuals. However, the perception that non-White populations might be less likely to test positive could stem from several factors related to systemic inequities, such as disparities in testing access and social determinants of health [50, 51, 53]. Also, according to Vandenberg [54], black people are more likely to live in densely populated, multi-generational housing, where social distancing can be difficult, further increasing the risk of transmission within households.

All the smooth terms for the probability of testing positive were statistically significant, supporting the assumption of nonlinearity for these covariates, and indicating that the increase in the goodness of fit outweighs the increased complexity of the model. Note that the approximate significance of the smooths represents a test that the indicated smooth is a flat or zero function, equivalent to the null hypothesis that a coefficient in a linear model or the variable has no effect. The results in Table 6 indicate strong evidence against the null hypothesis for each of the covariates, as reflected in the strong nonlinearity of the estimated smooths in Appendix 3. The reported effective degrees of freedom (EDF) in Table 6 quantify the complexity of a smooth function, essentially indicating how closely the smooth term follows a linear relationship or how wiggly it is. A higher EDF suggests a more complex, non-linear relationship (see the results in Appendix 3). The plots in Appendix 3 estimate the partial effects of each covariate on the link scale of the response variable, assuming all other predictor variables are zero and the smooths are centered at zero to preserve identifiability.

Specifically, and using the results in Table 6 and Appendix 3, the probability of testing positive at the ZCTAs level varies non-linearly across the total religious adherence rate per 1,000 individuals (TRATE_2020) gradient, with the highest levels of testing positive at lower levels of adherent rate (< 40), with a decreasing gradient. The probability of testing positive within ZCTAs is relatively lower and remains similar across adherence rates ranging from 40% to 80%. Notice a slight increase and greater variability in testing positive for COVID-19 at a rate above 80. The relationship between religious adherence and COVID-19 infection rates is not straightforward [54]. In general, research suggests a complex and sometimes contradictory relationship between religious adherence and testing positive for COVID-19. It depends on various factors, including the specific spiritual practices, beliefs, and the overall context of public health measures and community engagement. Religious gatherings and practices may increase the risk of exposure, and certain religious beliefs (such as the belief that God will protect one from illness) can lower adherence to public health recommendations (e.g., mask-wearing, vaccination). There is some evidence supporting a positive association between higher religious commitment and lower overall trust in science. However, religion can also play a positive role through religious leaders and communities, who can significantly promote and adhere to public health guidelines [55, 56, 57].

The plotted smooth curve for the social vulnerability index (SVI) is also wavy, with narrow variability from 0 to 0.78. It decreases sharply to 0.85 and then increases rapidly, with a noticeably high level of uncertainty in the estimate. These results suggest that the SVI between 0 and 0.78 index has limited positive and negative effects on testing positive for COVID-19. As the value of the SVI index increases, it first reduces the probability of testing positive, and it rapidly increases the likelihood at higher levels of the SVI. Various studies [58, 59, 60] conclude that areas with high social vulnerability, as indicated by factors such as poverty, inadequate transportation, and overcrowded housing, tend to experience more severe impacts from the pandemic.

The plot of the COVID-19 Vaccine Coverage Index (CVAC) variable is funnel-shaped (see the first part of Appendix 3). The wider sections of the funnel indicate areas where there is greater variability in the response for a given value of the predictor. The narrower sections of the funnel suggest that the variability of the response is less in those regions of the predictor variable’s range. The funnel is widening with a positive slope between 0 and 0.4 CVAC, and it becomes narrow and plateaus after 0.4 CVAC. The results suggest that the likelihood of testing positive is increasing, but becomes less predictable in ZCTAs with a lower level of CVAC (≤ 0.4). Conversely, there is increasing certainty in predicting a positive COVID-19 test in ZCTAs with a CVAC above 0.4. As explained above, there are five elements included when calculating CVAC: historic undervaccination, which provides for measures of standard vaccination coverage and vaccine exemption rate; sociodemographic barriers, aggregating variables measuring socioeconomically disadvantaged, and lack of access to information; resource-constrained healthcare system, combining measures healthcare workforce and strength of healthcare infrastructure; healthcare accessibility measured by combining underinsured, delayed care-seeking, lack of transport and limited connectivity variables, and irregular care seeking behavior that conjoin the lack of personal doctor and failure to seek routine care. These elements vary by ZCTAs and are unbalanced at lower CVAC. As the value of CVAC increases, these elements become balanced and their predictive power becomes certain.

Generally, the Cook’s partisan voter index (PVI) plot exhibits an upward trend below 30 and a downward trend thereafter. It is negative (below 0) and uncertain (high interval) for the PVI less than 0, implying a positive relationship between testing positive for COVID-19 in ZCTAs that lean Republican and democratic for PVI between 0 and 30, and a negative relationship between testing positive for COVID-19 in ZCTAs with PVI greater than 30. Therefore, testing positive for COVID-19 was more likely in ZCTAs that vote Republican and less likely in ZCTAs that vote Democratic. These results support the findings of Neelon et al. [61] and Roberts & Utych [62], which may be due to low vaccination rates among Republican voters [63, 64].

The smooth terms for the year and regions (STATEABB) represent QQ-plots for the factor variables describing the year of data correction (i.e., 2020, 2021, and 2022), and the 12 regions shown in Figure 1, respectively. The two variables are simply smooth with random effects in the model, thereby estimating random intercepts. The plotted Gaussian quantiles (x-axis) and random effects (y-axis) for both variables indicate that some years or regions had higher rates of COVID-19 positivity than others. The straight line shows that the random effects of the year and region variables are normally distributed (Gaussian) around testing positive for COVID-19, which supports the model assumption in terms of variable specification.

The last three plots in Table 6, which approximate the significance of smooth terms for testing positive, along with the two smooth plots of latitude and longitude, and a plot of the tensor product between latitude and longitude in Appendix 3, jointly model spatially varying effects across ZCTAs. The general results suggest that the spatial effect is non-linear, shifting based on location. The red regions in the last plot are where the interaction is positive (when the interaction has a more positive influence on testing positive). The yellow areas are when the interaction is negative (when the interaction has a more negative influence on testing positive). The ZCTAs with lower positivity rates were likely located between latitudes 30 and 34 and longitudes above -98, most likely in Oklahoma, Arkansas, and northern Louisiana.

The second part of Table 6 presents the results of Equation 2, which estimates the probability of dying after testing positive for COVID-19. The estimated parametric coefficients for age category, gender, and dummy variables representing Blacks and Hispanics were statistically significant at the 99 percent level, and the dummy variable for Whites was statistically significant at the 90 percent level. The results imply that there is a 1 or 10 percent less chance that the observed results occurred purely by random chance. The interpretation of the results is as explained above. The reference for the age category is more than 65 years. Compared to the reference age group (>60 years old), the probability of dying after testing positive decreased with decreasing age. It was lower by 1.65 percent compared to the 56-65 age category and lower by 7.00 percent, 14.99 percent, and 28.10 percent compared to the 40-55, 26-39, and ≤25 age categories, respectively. The results support the COVID-19-related deaths existing literature, indicating that young adults are not the most likely to die from COVID-19, and older adults and those with underlying health conditions are at higher risk of severe illness and death from COVID-19. For example, a study by Williamson et al. [16] concluded that COVID-19-related death was associated with greater age. The Centers for Disease Control and Prevention (CDC) also indicates that the risk of death increases significantly with advancing age, particularly for individuals over 65 years old. Over 81 percent of COVID-19 deaths occurred in people over age 65.

The results in Table 6 also indicate that male and black people were at higher risk (59.96% and 55.70%) of dying after testing positive compared to female and other races, respectively. Research shows that men were more likely to die from COVID-19 due to a combination of biological and behavioral factors. As reviewed by Fabião et al. [65], these variables include a potentially weaker immune response in men, genetic predispositions, unhealthy lifestyle choices, and a tendency for men to delay seeking medical care. Multiple factors contributed to higher COVID-19 mortality among Black individuals, reflecting the impact of systemic health disparities in the Southwest U.S. Notably, as Rogers et al. [66] show, black people are disproportionately represented in essential worker roles in industries like food service, transportation, and healthcare, increasing their risk of exposure to the virus. Also, limited access to testing and treatment, coupled with historical medical mistrust, may lead to delayed care and more severe disease outcomes [67]. The probability of dying after testing positive for COVID-19 among White and Hispanic people was lower by 47.79 and 42.32 percent, respectively, compared to other races. During the initial phases of the vaccination rollout, Black and Hispanic people were less likely than their white counterparts to be vaccinated [68, 69], which narrowed with time. It is crucial to understand that the pandemic exposed and exacerbated existing health and social inequities, impacting different racial and ethnic groups in varied circumstances [70, 71].

The last part of Table 6 shows the approximate significance of smooth terms for the probability of dying after testing positive for COVID-19. From the table, the variables measuring the social vulnerability index (SVI), Vaccine Coverage Index (CVAC), and the national workability index (NatWalkInd) were statistically non-significant; see also their plots in Appendix 3. The results in Table 6 and Appendix 3 also show that the probability of dying after testing positive varies non-linearly along the total religious adherence rate (TRATE_2020) gradient. The shape of the gradient implies that at an adherence rate of less than 60, the probability of dying increases, then starts decreasing thereafter. Because the predicted line is below zero, the overall effect of religiosity is negative; that is, religiosity decreases the likelihood of dying after testing positive for COVID-19. Some studies [72, 73] have investigated the relationship between religiosity and COVID-19 outcomes, suggesting that religious coping mechanisms and social support within religious communities may play a role in mitigating the pandemic’s negative impacts in ZCTAs with a high rate of religious adherence.

Regarding the Cook’s partisan voter index (PVI) smooth plot, it exhibits a downward trend from positive values of the response link to negative values, implying the probability of dying after testing positive for COVID-19 is higher in ZCTAs that vote Republican and very low in ZCTAs that vote Democratic. However, it is crucial to understand that political affiliation is not a direct cause of higher mortality. Political affiliation influenced attitudes towards mitigation strategies, such as masking, social distancing, and vaccine uptake, which in turn affected the pandemic’s trajectory and impact [74]. The smooth plot of the Smart Location Index (SLC_score), which ranges in value from 0 (least location-efficient site, high congestion and pollution) to 100 (most location-efficient site, lower congestion and pollution), shows a significant decrease in the probability of dying for a score greater than 60 percent. The interplay of high pollution and congestion creates a synergistic effect, further increasing the risk of severe illness and death from COVID-19. However, while the links between pollution and COVID-19 mortality are well-established, more research is needed to fully understand the specific mechanisms involved and the impact of walkable, bikeable, and transit-friendly lifestyles on different populations [75].

The ZCTAs with high prevalence of Chronic Obstructive Pulmonary Disease (propCOPD) and obesity (propOBESITY) were more likely to experience high death rates of people who tested positive for COVID-19. The probability of dying increases continuously with the former, and it increases but then plateaus at 45 percent with the latter. Both Chronic Obstructive Pulmonary Disease (COPD) and obesity are significant risk factors for worse outcomes in COVID-19 patients, including increased mortality [76, 77]. While COPD diminishes lung function, making it harder to cope with the damage inflicted by the virus, excess weight can exacerbate breathing difficulties, particularly when dealing with a respiratory infection like COVID-19 [78]. The last three plots in Appendix 3 illustrate how the predicted probability of dying changes across the geographical area (ZCTAs) represented by latitude and longitude coordinates, while holding all other variables in the model at their mean. The plot’s color or contour lines illustrate the magnitude of this effect. Red colors indicate areas with higher predicted response values, and green colors suggest lower predicted values. By combining the results of the last three plots, the probability of dying increases from the southern to the northern regions of the study area. The primary differences between Southern and Northern regions lie in their geography, culture, and economy. For example, Southern Louisiana is known for its marshes, bayous, and Cajun culture, while Northern Louisiana features rolling hills, pine forests, and a more traditional Southern culture. While Southern Louisiana has a strong French influence, particularly in the Cajun and Creole cultures, Northern Louisiana exhibits a more traditional Southern culture, with influences from Texas and neighboring states. Culture has a significant impact on health beliefs and practices, affecting everything from how people perceive illness to their approaches to healthcare and health-seeking behaviors [79, 80, 81].

## SUMMARY AND IMPLICATIONS FOR PUBLIC POLICY

The COVID-19 pandemic significantly impacted global health, with millions infected and a substantial number of deaths reported worldwide. The virus, SARS-CoV-2, has been associated with diverse outcomes, ranging from asymptomatic infections to severe illness and death. Consequently, various researchers have studied the causal relationship between regional and meso-level factors and the likelihood of testing positive for or dying from the virus. While regional variables have broader implications for public health policy, individual-level analysis helps explain a person’s traits or behaviors, which is vital in identifying the characteristics of the human decision-making process and group health outcomes. The current extant literature on COVID-19 lacks studies that use extensive data and jointly analyze factors associated with testing positive and subsequent COVID-19-related deaths. Moreover, the majority of studies use small samples and focus on either testing positive or death.

This study utilizes data from the Louisiana Department of Health and Hospitals, obtained through a special request, in conjunction with meso-level variables from various Geographical Information Systems and Platforms. For each reported COVID-19 case, the patient has a unique identification number and demographic characteristics, including age, gender, race, zip code, and the date the patient tested positive or died. The meso-variables included measures of social vulnerability, vaccine distribution challenges, political and religious beliefs, indicators related to the built environment, and location efficiency of various areas in the United States. Data analysis involved jointly estimating the probability of testing positive and dying from the virus using a spatial model within a generalized additive models framework. The model establishes the spatial relationships between covariates and the two response variables through smooth patterns. The relationship can be linear or non-linear, thereby reducing spatial heterogeneity and minimizing the effects of autocorrelation among predictor variables.

The main results suggest a strong real effect between testing positive and individual-level covariates. Particularly, the probability of testing positive increases with age when compared to individuals older than 65. Black, White, and Hispanic people, compared to other races, were likely to test positive for COVID-19. The probability of testing positive at the ZCTA level varied non-linearly across the total religious adherence rate, social vulnerability index, the Cook’s partisan voter index, and the spatial effect is non-linear, shifting based on location. Regarding estimating the probability of dying after testing positive for COVID-19, the individual-level variables such as age, gender, and dummy variables representing Blacks and Hispanics were statistically significant. The variables measuring the social vulnerability, vaccine coverage, and the national workability index were statistically non-significant. The probability of dying after testing positive varies non-linearly with measures of religiosity, political partisanship, and congestion and pollution. The areas with high prevalence of Chronic Obstructive Pulmonary Disease and obesity were more likely to experience high death rates, with the predicted probability of dying increasing from the southern to the northern regions of the study area.

To mitigate the impact of future pandemics like COVID-19, public health policies should specifically focus on addressing existing health disparities. There must be data-driven approaches and an expansion of the evidence base by collecting and analyzing health data disaggregated by age, race, and ethnicity to identify and quantify disparities in real-time during a pandemic response. Granular data enables the efficient allocation of resources to vulnerable groups within the population, facilitating a deeper understanding of disparities and improving targeted interventions. There is also a need to develop new or refine existing methods for systematically tracking these disparities, and incorporating social vulnerability markers into data collection and analysis to understand the influence of social and economic factors on health outcomes across different age and racial/ethnic groups.

Fostering meaningful engagement with community institutions and diverse leaders, especially in historically marginalized communities, to understand their needs and develop culturally competent interventions. Collaboration with community and faith-based groups would tailor interventions and address community priorities. Based on the results of this study, future pandemics require developing health communication strategies that address distrust in healthcare institutions, which is often prevalent in communities of color due to historical mistreatment. Utilizing trusted community leaders and health workers to disseminate public health messaging would encourage adherence to prevention measures and the uptake of interventions, such as testing and vaccinations. Moreover, collaborating with community organizations and leveraging community health workers to identify and address health-related social needs that may disproportionately impact specific age and racial/ethnic groups during a pandemic.

The region needs to address disparities in access to care by investing in telemedicine, expanding healthcare infrastructure in underserved communities, and supporting mobile health services to reach populations in rural areas and those facing barriers to traditional healthcare settings. There is also a demand to recognize that inequities in social determinants of health drive health disparities and must be addressed through intersectoral policies that promote health equity and racial justice. Implementing policies that improve access to robust built environments, which minimize congestion and pollution, and creating a strong public health infrastructure are additional methods to prepare for future pandemics. Investing in pandemic preparedness in low-resource settings is vital to prevent future outbreaks from disproportionately affecting these areas. By implementing targeted public health policies that focus on determinants of health disparities, governments can better prepare for and respond to future pandemics more equitably, thereby reducing health disparities and protecting the health and well-being of all populations. Proven and scientific public health considerations should guide future policy decisions, minimizing the influence of political ideologies and culture.

## Data Availability

Some data sources are contained in the manuscrispt.

---

**Appendix 1.**
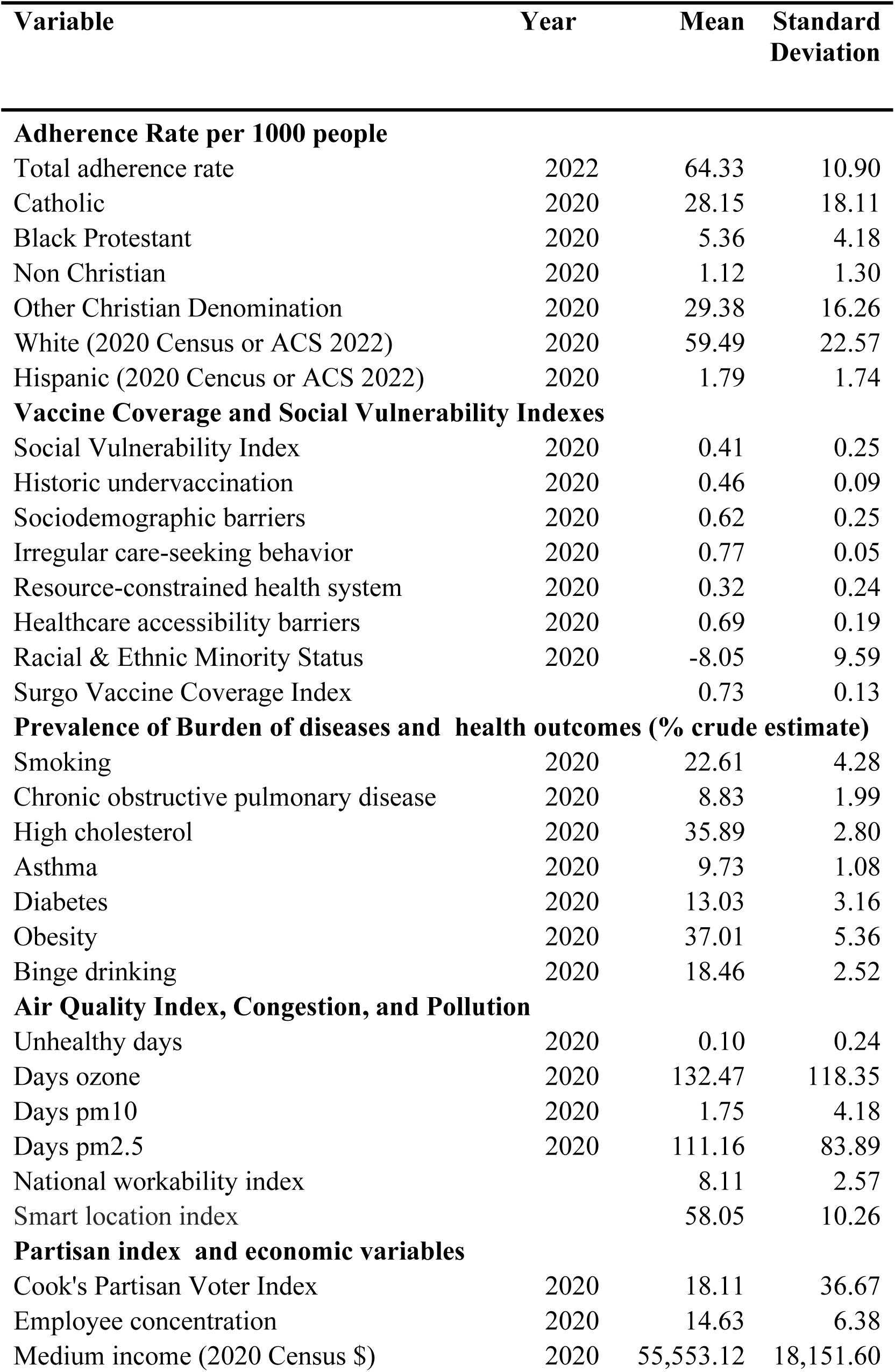
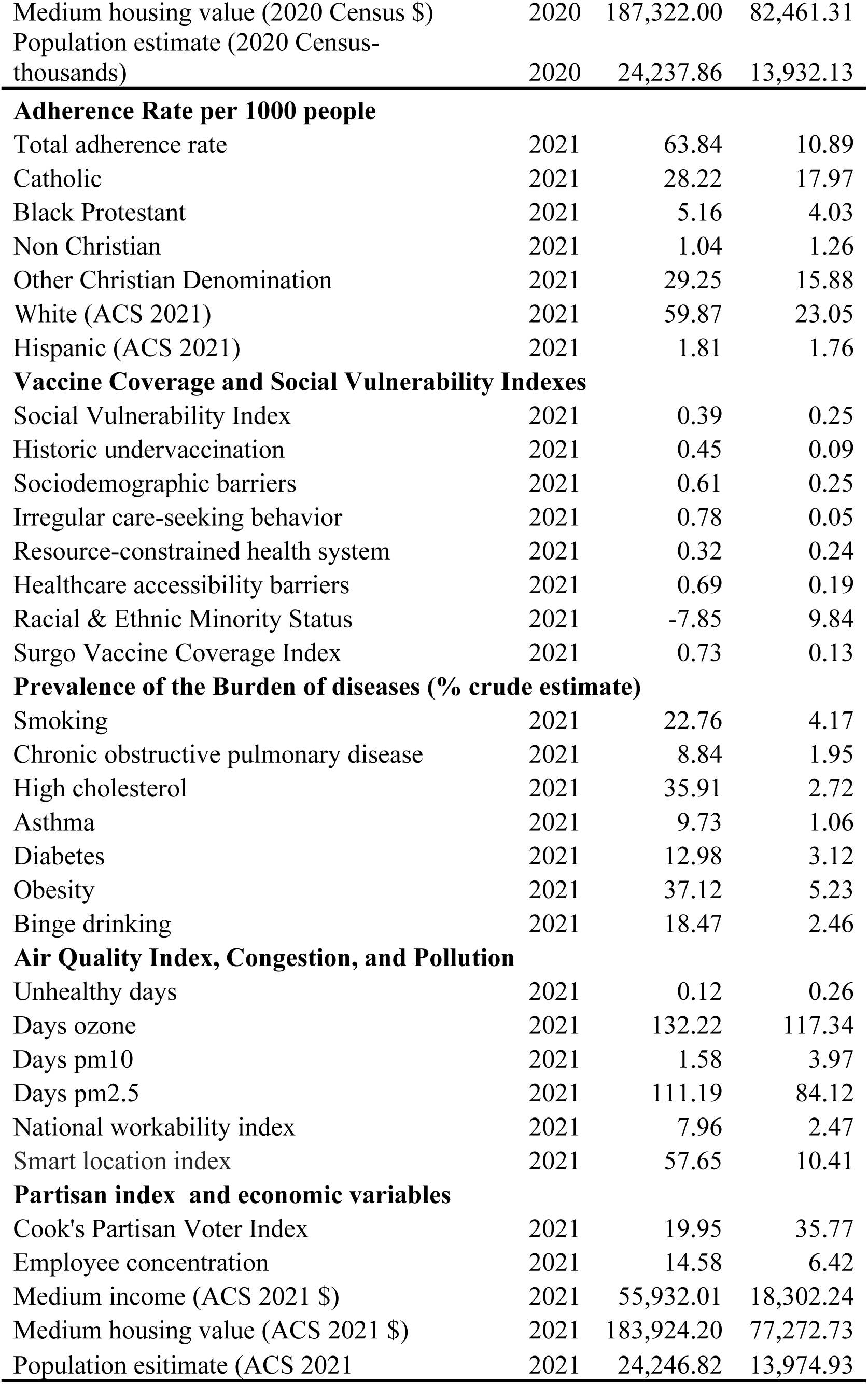
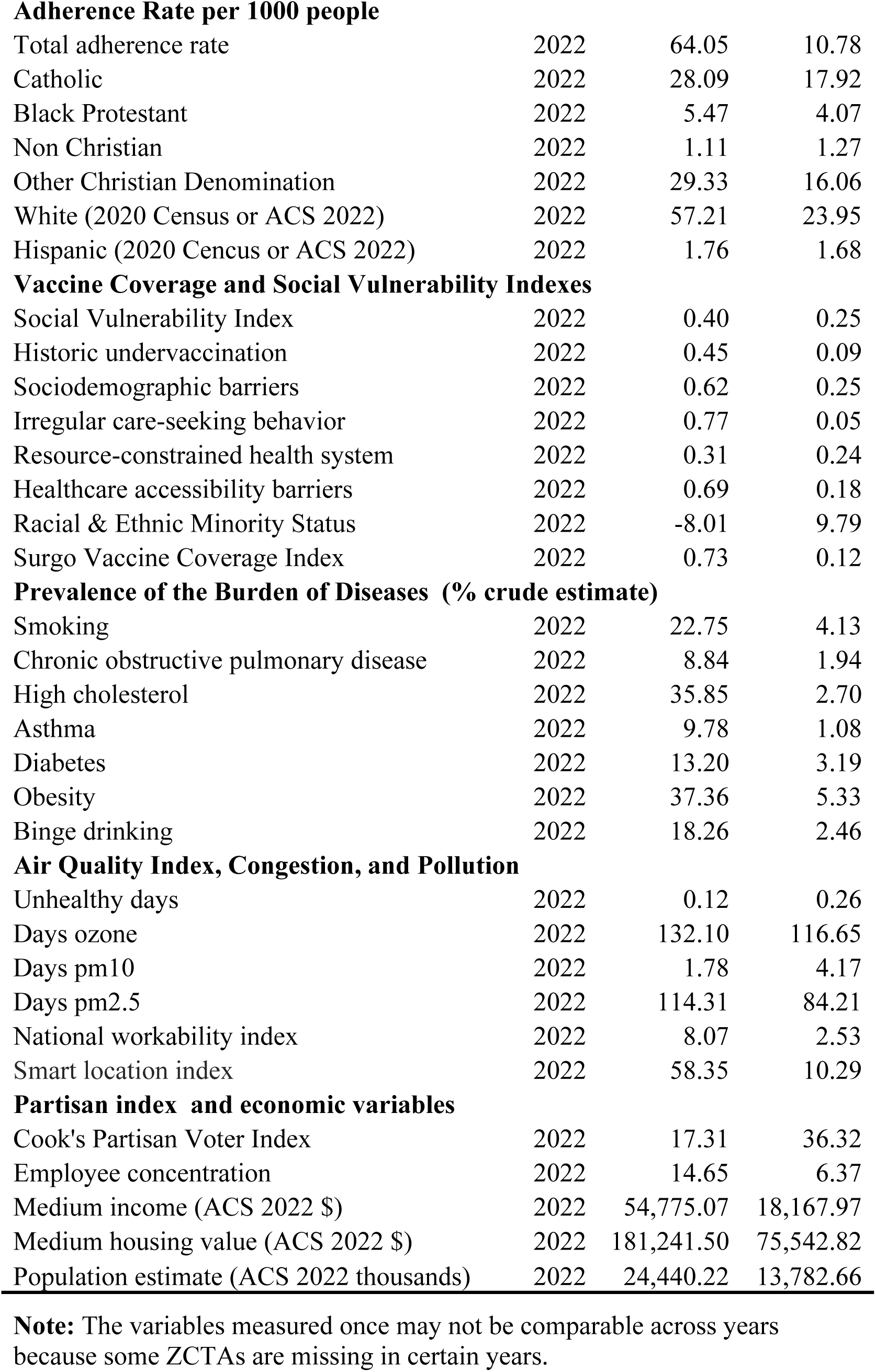
The list of variables included in the global baseline model.

| Variable | Year | Mean | Standard<br>Deviation |
| --- | --- | --- | --- |
| **Adherence Rate per 1000 people** |  |  |  |
| Total adherence rate | 2022 | 64.33 | 10.90 |
| Catholic | 2020 | 28.15 | 18.11 |
| Black Protestant | 2020 | 5.36 | 4.18 |
| Non Christian | 2020 | 1.12 | 1.30 |
| Other Christian Denomination | 2020 | 29.38 | 16.26 |
| White (2020 Census or ACS 2022) | 2020 | 59.49 | 22.57 |
| Hispanic (2020 Cencus or ACS 2022) | 2020 | 1.79 | 1.74 |
| **Vaccine Coverage and Social Vulnerability Indexes** |  |  |  |
| Social Vulnerability Index | 2020 | 0.41 | 0.25 |
| Historic undervaccination | 2020 | 0.46 | 0.09 |
| Sociodemographic barriers | 2020 | 0.62 | 0.25 |
| Irregular care-seeking behavior | 2020 | 0.77 | 0.05 |
| Resource-constrained health system | 2020 | 0.32 | 0.24 |
| Healthcare accessibility barriers | 2020 | 0.69 | 0.19 |
| Racial & Ethnic Minority Status | 2020 | -8.05 | 9.59 |
| Surgo Vaccine Coverage Index |  | 0.73 | 0.13 |
| **Prevalence of Burden of diseases and health outcomes (% crude estimate)** |  |  |  |
| Smoking | 2020 | 22.61 | 4.28 |
| Chronic obstructive pulmonary disease | 2020 | 8.83 | 1.99 |
| High cholesterol | 2020 | 35.89 | 2.80 |
| Asthma | 2020 | 9.73 | 1.08 |
| Diabetes | 2020 | 13.03 | 3.16 |
| Obesity | 2020 | 37.01 | 5.36 |
| Binge drinking | 2020 | 18.46 | 2.52 |
| **Air Quality Index, Congestion, and Pollution** |  |  |  |
| Unhealthy days | 2020 | 0.10 | 0.24 |
| Days ozone | 2020 | 132.47 | 118.35 |
| Days pm10 | 2020 | 1.75 | 4.18 |
| Days pm2.5 | 2020 | 111.16 | 83.89 |
| National workability index |  | 8.11 | 2.57 |
| Smart location index |  | 58.05 | 10.26 |
| **Partisan index and economic variables** |  |  |  |
| Cook's Partisan Voter Index | 2020 | 18.11 | 36.67 |
| Employee concentration | 2020 | 14.63 | 6.38 |
| Medium income (2020 Census \$) | 2020 | 55,553.12 | 18,151.60 |
| Medium housing value (2020 Census \$) | 2020 | 187,322.00 | 82,461.31 |
| Population estimate (2020 Census-thousands) | 2020 | 24,237.86 | 13,932.13 |
| **Adherence Rate per 1000 people** |  |  |  |
| Total adherence rate | 2021 | 63.84 | 10.89 |
| Catholic | 2021 | 28.22 | 17.97 |
| Black Protestant | 2021 | 5.16 | 4.03 |
| Non Christian | 2021 | 1.04 | 1.26 |
| Other Christian Denomination | 2021 | 29.25 | 15.88 |
| White (ACS 2021) | 2021 | 59.87 | 23.05 |
| Hispanic (ACS 2021) | 2021 | 1.81 | 1.76 |
| **Vaccine Coverage and Social Vulnerability Indexes** |  |  |  |
| Social Vulnerability Index | 2021 | 0.39 | 0.25 |
| Historic undervaccination | 2021 | 0.45 | 0.09 |
| Sociodemographic barriers | 2021 | 0.61 | 0.25 |
| Irregular care-seeking behavior | 2021 | 0.78 | 0.05 |
| Resource-constrained health system | 2021 | 0.32 | 0.24 |
| Healthcare accessibility barriers | 2021 | 0.69 | 0.19 |
| Racial & Ethnic Minority Status | 2021 | -7.85 | 9.84 |
| Surgo Vaccine Coverage Index | 2021 | 0.73 | 0.13 |
| **Prevalence of the Burden of diseases (% crude estimate)** |  |  |  |
| Smoking | 2021 | 22.76 | 4.17 |
| Chronic obstructive pulmonary disease | 2021 | 8.84 | 1.95 |
| High cholesterol | 2021 | 35.91 | 2.72 |
| Asthma | 2021 | 9.73 | 1.06 |
| Diabetes | 2021 | 12.98 | 3.12 |
| Obesity | 2021 | 37.12 | 5.23 |
| Binge drinking | 2021 | 18.47 | 2.46 |
| **Air Quality Index, Congestion, and Pollution** |  |  |  |
| Unhealthy days | 2021 | 0.12 | 0.26 |
| Days ozone | 2021 | 132.22 | 117.34 |
| Days pm10 | 2021 | 1.58 | 3.97 |
| Days pm2.5 | 2021 | 111.19 | 84.12 |
| National workability index | 2021 | 7.96 | 2.47 |
| Smart location index | 2021 | 57.65 | 10.41 |
| **Partisan index and economic variables** |  |  |  |
| Cook's Partisan Voter Index | 2021 | 19.95 | 35.77 |
| Employee concentration | 2021 | 14.58 | 6.42 |
| Medium income (ACS 2021 \$) | 2021 | 55,932.01 | 18,302.24 |
| Medium housing value (ACS 2021 \$) | 2021 | 183,924.20 | 77,272.73 |
| Population estimate (ACS 2021) | 2021 | 24,246.82 | 13,974.93 |
| **Adherence Rate per 1000 people** |  |  |  |
| Total adherence rate | 2022 | 64.05 | 10.78 |
| Catholic | 2022 | 28.09 | 17.92 |
| Black Protestant | 2022 | 5.47 | 4.07 |
| Non Christian | 2022 | 1.11 | 1.27 |
| Other Christian Denomination | 2022 | 29.33 | 16.06 |
| White (2020 Census or ACS 2022) | 2022 | 57.21 | 23.95 |
| Hispanic (2020 Census or ACS 2022) | 2022 | 1.76 | 1.68 |
| **Vaccine Coverage and Social Vulnerability Indexes** |  |  |  |
| Social Vulnerability Index | 2022 | 0.40 | 0.25 |
| Historic undervaccination | 2022 | 0.45 | 0.09 |
| Sociodemographic barriers | 2022 | 0.62 | 0.25 |
| Irregular care-seeking behavior | 2022 | 0.77 | 0.05 |
| Resource-constrained health system | 2022 | 0.31 | 0.24 |
| Healthcare accessibility barriers | 2022 | 0.69 | 0.18 |
| Racial & Ethnic Minority Status | 2022 | -8.01 | 9.79 |
| Surgo Vaccine Coverage Index | 2022 | 0.73 | 0.12 |
| **Prevalence of the Burden of Diseases (% crude estimate)** |  |  |  |
| Smoking | 2022 | 22.75 | 4.13 |
| Chronic obstructive pulmonary disease | 2022 | 8.84 | 1.94 |
| High cholesterol | 2022 | 35.85 | 2.70 |
| Asthma | 2022 | 9.78 | 1.08 |
| Diabetes | 2022 | 13.20 | 3.19 |
| Obesity | 2022 | 37.36 | 5.33 |
| Binge drinking | 2022 | 18.26 | 2.46 |
| **Air Quality Index, Congestion, and Pollution** |  |  |  |
| Unhealthy days | 2022 | 0.12 | 0.26 |
| Days ozone | 2022 | 132.10 | 116.65 |
| Days pm10 | 2022 | 1.78 | 4.17 |
| Days pm2.5 | 2022 | 114.31 | 84.21 |
| National workability index | 2022 | 8.07 | 2.53 |
| Smart location index | 2022 | 58.35 | 10.29 |
| **Partisan index and economic variables** |  |  |  |
| Cook's Partisan Voter Index | 2022 | 17.31 | 36.32 |
| Employee concentration | 2022 | 14.65 | 6.37 |
| Medium income (ACS 2022 \$) | 2022 | 54,775.07 | 18,167.97 |
| Medium housing value (ACS 2022 \$) | 2022 | 181,241.50 | 75,542.82 |
| Population estimate (ACS 2022 thousands) | 2022 | 24,440.22 | 13,782.66 |

**Note:** The variables measured once may not be comparable across years because some ZCTAs are missing in certain years.

**Appendix 2.**
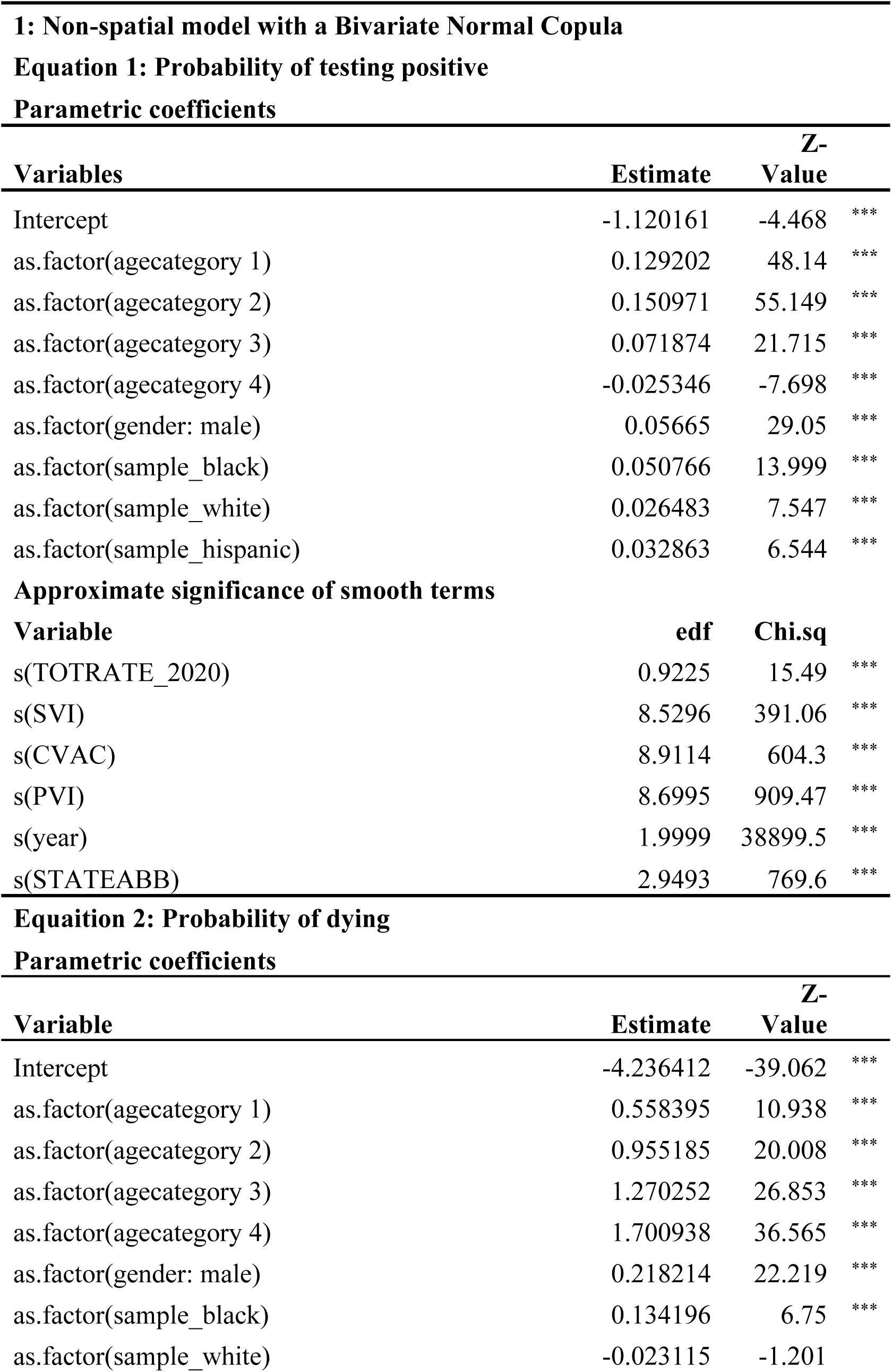
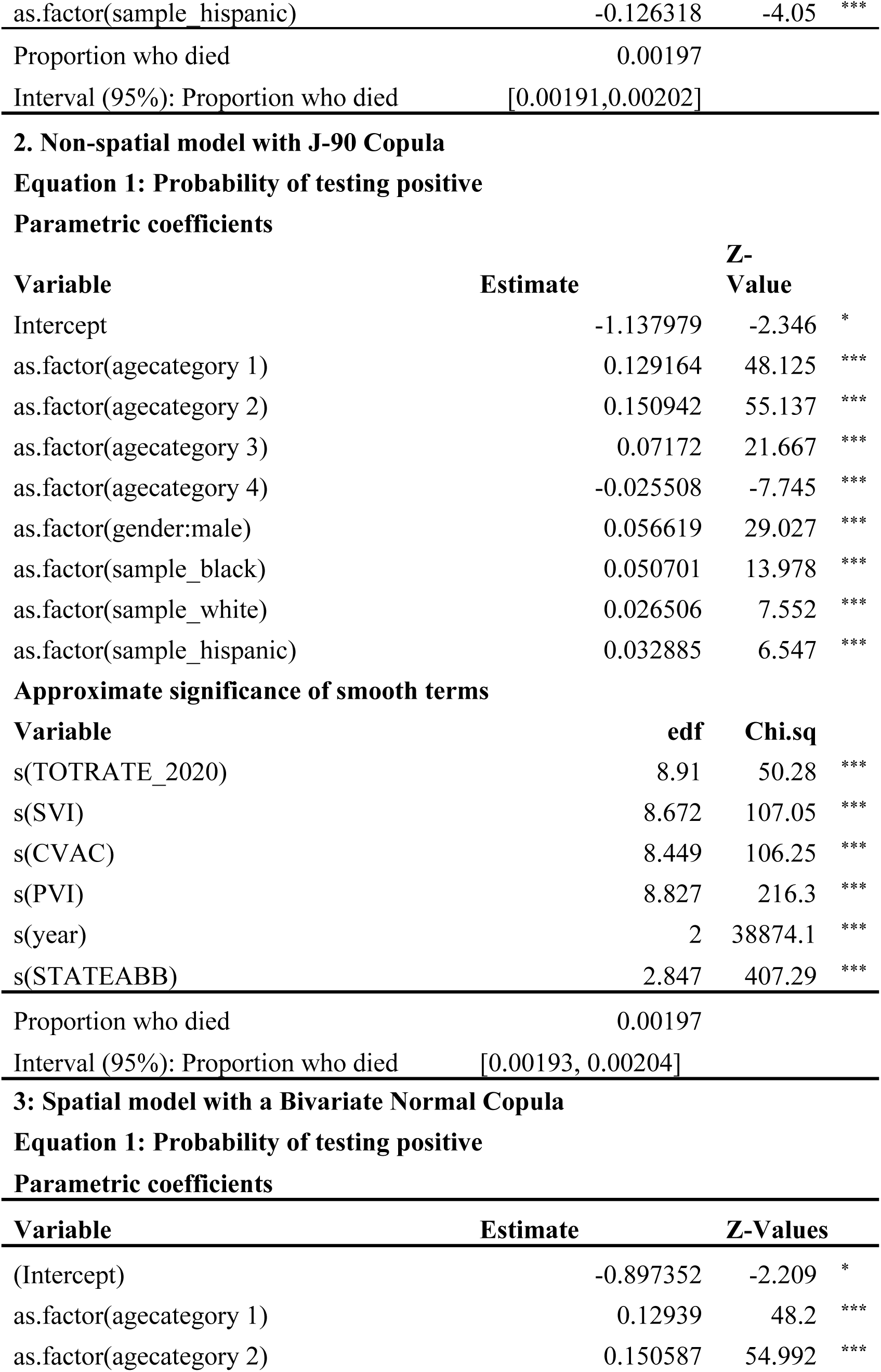
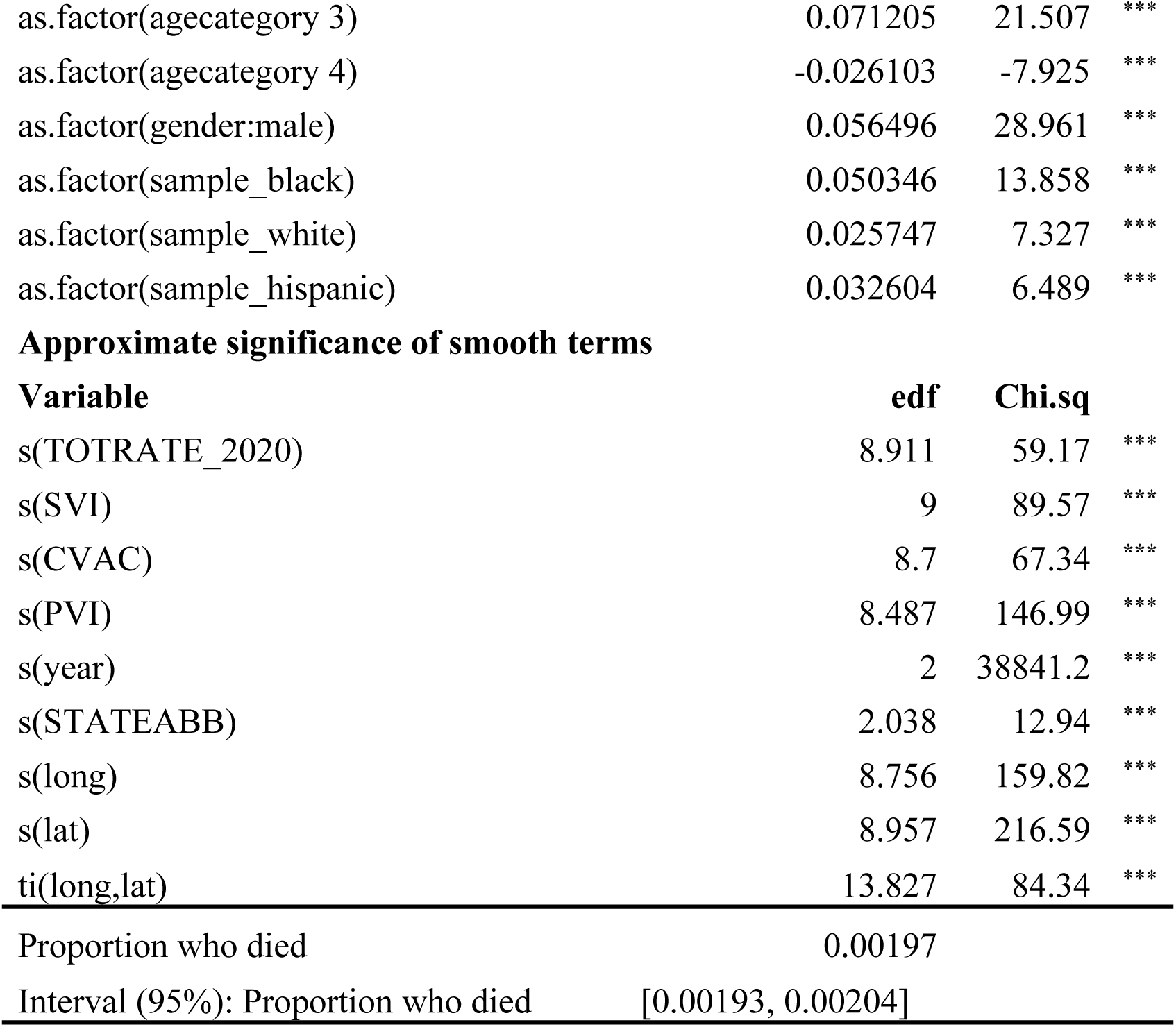
Results of the tested generalized additive models.

Parametric coefficients
| Variables | Estimate | Z-Value |  |
| --- | --- | --- | --- |
| Intercept | -1.120161 | -4.468 | *** |
| as.factor(agecategory 1) | 0.129202 | 48.14 | *** |
| as.factor(agecategory 2) | 0.150971 | 55.149 | *** |
| as.factor(agecategory 3) | 0.071874 | 21.715 | *** |
| as.factor(agecategory 4) | -0.025346 | -7.698 | *** |
| as.factor(gender: male) | 0.05665 | 29.05 | *** |
| as.factor(sample_black) | 0.050766 | 13.999 | *** |
| as.factor(sample_white) | 0.026483 | 7.547 | *** |
| as.factor(sample_hispanic) | 0.032863 | 6.544 | *** |

Approximate significance of smooth terms
| Variable | edf | Chi.sq |  |
| --- | --- | --- | --- |
| s(TOTRATE_2020) | 0.9225 | 15.49 | *** |
| s(SVI) | 8.5296 | 391.06 | *** |
| s(CVAC) | 8.9114 | 604.3 | *** |
| s(PVI) | 8.6995 | 909.47 | *** |
| s(year) | 1.9999 | 38899.5 | *** |
| s(STATEABB) | 2.9493 | 769.6 | *** |

Equation 2: Probability of dying Parametric coefficients
| Variable | Estimate | Z-Value |  |
| --- | --- | --- | --- |
| Intercept | -4.236412 | -39.062 | *** |
| as.factor(agecategory 1) | 0.558395 | 10.938 | *** |
| as.factor(agecategory 2) | 0.955185 | 20.008 | *** |
| as.factor(agecategory 3) | 1.270252 | 26.853 | *** |
| as.factor(agecategory 4) | 1.700938 | 36.565 | *** |
| as.factor(gender: male) | 0.218214 | 22.219 | *** |
| as.factor(sample_black) | 0.134196 | 6.75 | *** |
| as.factor(sample_white) | -0.023115 | -1.201 |  |
| as.factor(sample_hispanic) | -0.126318 | -4.05 | *** |
| Proportion who died | 0.00197 |  |  |
| Interval (95%): Proportion who died | [0.00191,0.00202] |  |  |

Parametric coefficients
| Variable | Estimate | Z-Value |  |
| --- | --- | --- | --- |
| Intercept | -1.137979 | -2.346 | * |
| as.factor(agecategory 1) | 0.129164 | 48.125 | *** |
| as.factor(agecategory 2) | 0.150942 | 55.137 | *** |
| as.factor(agecategory 3) | 0.07172 | 21.667 | *** |
| as.factor(agecategory 4) | -0.025508 | -7.745 | *** |
| as.factor(gender:male) | 0.056619 | 29.027 | *** |
| as.factor(sample_black) | 0.050701 | 13.978 | *** |
| as.factor(sample_white) | 0.026506 | 7.552 | *** |
| as.factor(sample_hispanic) | 0.032885 | 6.547 | *** |

Approximate significance of smooth terms
| Variable | edf | Chi.sq |  |
| --- | --- | --- | --- |
| s(TOTRATE_2020) | 8.91 | 50.28 | *** |
| s(SVI) | 8.672 | 107.05 | *** |
| s(CVAC) | 8.449 | 106.25 | *** |
| s(PVI) | 8.827 | 216.3 | *** |
| s(year) | 2 | 38874.1 | *** |
| s(STATEABB) | 2.847 | 407.29 | *** |
| Proportion who died | 0.00197 |  |  |
| Interval (95%): Proportion who died | [0.00193, 0.00204] |  |  |

Parametric coefficients
| Variable | Estimate | Z-Values |  |
| --- | --- | --- | --- |
| (Intercept) | -0.897352 | -2.209 | * |
| as.factor(agecategory 1) | 0.12939 | 48.2 | *** |
| as.factor(agecategory 2) | 0.150587 | 54.992 | *** |
| as.factor(agecategory 3) | 0.071205 | 21.507 | *** |
| as.factor(agecategory 4) | -0.026103 | -7.925 | *** |
| as.factor(gender:male) | 0.056496 | 28.961 | *** |
| as.factor(sample_black) | 0.050346 | 13.858 | *** |
| as.factor(sample_white) | 0.025747 | 7.327 | *** |
| as.factor(sample_hispanic) | 0.032604 | 6.489 | *** |

Approximate significance of smooth terms
| Variable | edf | Chi.sq |  |
| --- | --- | --- | --- |
| s(TOTRATE_2020) | 8.911 | 59.17 | *** |
| s(SVI) | 9 | 89.57 | *** |
| s(CVAC) | 8.7 | 67.34 | *** |
| s(PVI) | 8.487 | 146.99 | *** |
| s(year) | 2 | 38841.2 | *** |
| s(STATEABB) | 2.038 | 12.94 | *** |
| s(long) | 8.756 | 159.82 | *** |
| s(lat) | 8.957 | 216.59 | *** |
| ti(long,lat) | 13.827 | 84.34 | *** |

|  |  |
| --- | --- |
| Proportion who died | 0.00197 |
| Interval (95%): Proportion who died | [0.00193, 0.00204] |

**Appendix 3.**
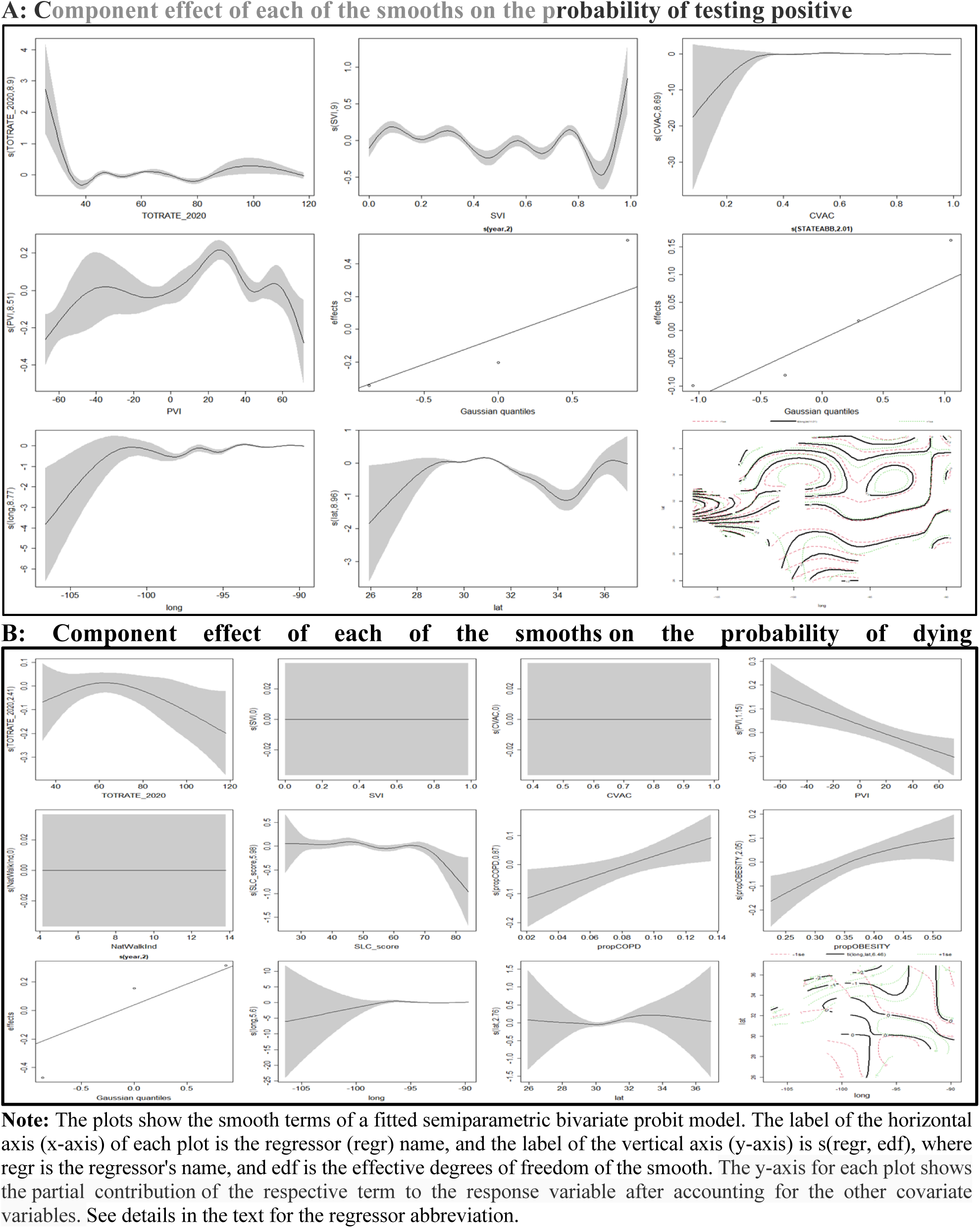
Plot of partial effects of some predictors with a 95 percent confidence interval A: Component effect of each of the smooths on the probability of testing positive B: Component effect of each of the smooths on the probability of dying.

## Notes

### Competing Interest Statement

The authors have declared no competing interest.

### Funding Statement

The author(s) received no specific funding for this work.

### Author Declarations

Louisiana Department of Health, Office of Public Health. The data was de-identified by the Louisiana Department of Health before approval.

